# Food additive exposure associated with reduction in gut microbiota diversity

**DOI:** 10.64898/2026.06.22.26356234

**Authors:** Rohan Singh, Daniel McDonald, Rob Knight, Marcel Salathé

## Abstract

Consumption of ultra-processed foods is rising globally and has been implicated in inflammation and metabolic dysfunction, yet the impact of specific food additives on the human gut microbiota remains poorly understood. Using dietary data from the Food & You study (∼1000 participants in Switzerland), we identified 257 distinct additives from 4,119 unique packaged products to quantify each participant’s daily additive exposure. Higher exposure to a combination of high intensity sweeteners and sugar polyols, commonly found in low calorie products, was independently associated with reduced gut microbial Shannon diversity (β = −0.39, p < 0.001), after adjustment for demographics, diet quality, BMI and bowel movement frequency. At a broader level, total additive exposure and fast food consumption were each negatively associated with gut microbial diversity; however, additive exposure remained independently associated and also specifically attenuated the diversity benefits of vegetable rich diets. Furthermore, microbial log ratio signatures linked to additive exposure showed strong negative correlations with Shannon diversity, including emulsifiers and thickeners (r = −0.66) and preservatives and antioxidants (r = −0.56). Integrating additive exposure with healthy dietary components such as HEI, fruits, or vegetables strengthened associations with gut microbial diversity; for example, vegetable linked correlations with Shannon diversity increased from r = 0.52 to r = 0.65 when contrasted against preservative-antioxidant exposure. Concordantly, microbial signatures associated with the sweeteners and sugar polyols additive combination showed depletion of fiber associated commensal taxa, and enrichment of pathways involved in polyol and aromatic compound metabolism. Notably, these associations emerged despite packaged foods representing only ∼15% of logged dietary intake, underscoring the sensitivity of gut microbial diversity to limited exposure, and demonstrating that without integrating additive and processed-food metrics, one of the largest effect-size phenomena in human gut microbiota diversity would remain undetected.

## Introduction

Modern dietary patterns show a concerning global trend toward ultra-processed foods (UPFs), industrial formulations of refined ingredients, additives, and synthetic constituents that comprise over 50% of total energy intake in high-income nations such as USA, Canada and the UK, and are rapidly gaining prominence (20–33% of dietary energy) in middle-income countries^1^. Epidemiological data link high UPF consumption to elevated risks of inflammatory bowel disease, colorectal cancer and metabolic syndrome^2–6^, although these cross-sectional cohort studies have methodological limitations that prevent causal inference. Most rely on food frequency questionnaires (FFQs) not designed to capture ingredient-level details, making it challenging to distinguish the potential effects of individual additives versus their combined presence in UPFs.

Experimental evidence points to several mechanisms through which food additives may disrupt gut microbial ecosystems. Synthetic emulsifiers such as carboxymethylcellulose (CMC) and polysorbate-80 (P80) perturb gut microbial communities in mouse models, promoting inflammation and increasing metabolic disorder susceptibility^7,8^, and direct ex vivo exposure of human microbiota to twenty common emulsifiers showed compound-specific effects, with P80, maltodextrin, xanthan gum, and various carrageenans driving significant dysbiotic shifts and increased pro-inflammatory potential, while soy lecithin and mono- and diglycerides showed no impact^9^. Carrageenan isomers have been shown to deplete the anti-inflammatory bacterium Akkermansia muciniphila^10^, lower short-chain fatty acid levels, and thin the colonic mucus layer^11^, collectively priming the gut epithelial barrier for inflammation. Non-nutritive sweeteners can also alter gut microbiome composition and affect glucose metabolism in humans, with effects that vary across sweetener types and individuals, as demonstrated through controlled feeding and gnotobiotic transplantation experiments^12^. Sugar polyols such as sorbitol and maltitol, which are incompletely absorbed in the small intestine, can further alter microbial composition through osmotic and fermentative effects in a dose-dependent manner^13,14^.

Despite this experimental evidence, the specific contributions of individual additives to gut microbiota disruption in free-living human populations remain poorly understood. Most epidemiological studies of UPFs rely on broad NOVA classifications that do not distinguish between additive types or quantities, and although recent research has mapped additive co-occurrence patterns in the French population^15^, the aggregate impacts of real-world additive exposure on gut microbiota diversity have yet to be systematically tested.

To address the gap in population-level data on real-world food additive consumption and gut microbiota diversity patterns, we leveraged the Food & You digital cohort^18^, a diet- and glucose-tracking study of about 1,000 healthy Swiss adults. Participants documented their daily food intake over a 2–4 week period using a smartphone application (MyFoodRepo), with ∼15% of the recorded meals consisting of barcode-scanned packaged foods and beverages. By connecting 12,110 unique barcoded items to the OpenFoodFacts database^19^, we were able to construct profiles of individual additive exposures by calculating daily counts of distinct E-codes (the standardized numerical identifiers assigned to approved food additives under European food safety regulations), while simultaneously capturing temporal patterns, demographic factors, and overall dietary quality metrics.

We employed non-negative matrix factorization to identify naturally occurring patterns of additive co-occurrence, revealing three predominant mixtures in the population. We examined how these exposure patterns relate to gut microbial ecosystems, and developed composite metrics that contrast dietary quality indicators against additive burden (e.g., HEI-minus-additives, vegetables-minus-sweeteners), enabling assessment of the relative importance of both beneficial dietary components and potentially detrimental additives in shaping gut microbiota diversity. Primarily, we aimed to (1) characterize real-world food additive exposure patterns and how they vary across dietary and demographic factors, (2) identify naturally co-occurring additive mixtures and assess their independent associations with gut microbial diversity, (3) testing whether additive exposure attenuates the microbiome benefits associated with protective dietary components, and (4) identify microbial taxa and predicted functional pathways associated with additive exposure. Together, these analyses provide insights that could inform more nuanced dietary recommendations incorporating both nutritional quality and additive exposure considerations.

## Results

### Acidity Regulators and Emulsifiers Dominate Exposure and Food Product Profiles

From the 12,110 unique barcoded foods consumed in the Food & You cohort, 4,119 (34%) contained at least one additive, with 257 distinct additives identified across all products. Additives were categorized according to their E-code designations and classified into: 1) colorants, 2) preservatives and antioxidants (PA), 3) thickeners, stabilizers, emulsifiers, and gelling agents (TSEGs), 4) pH regulators, anti-caking, and leavening agents (pH/AC/L), 5) flavor enhancers, 6) sweeteners and glazing agents (SG).

Among the additive categories, pH/AC/L constituted the largest additive group (Figure 1a) and also showed the highest participant exposure (Figure 1b, Supplementary Table 1), where prevalence refers to the percentage of participants exposed to the additive throughout the study period. This exposure pattern was driven primarily by citric acid (E330), which was present in 77% of participants, while carbon dioxide (E290), sodium citrate (E331), ammonium carbonates (E503), and sodium hydrogen carbonate (E500ii) were prevalent between 34–40%. TSEGs formed the second largest category with similarly widespread exposure, led by lecithins (E322i, E322) at 65–74%, and a range of additives including mono- and diglycerides of fatty acids (E471), modified starches (E14xx), xanthan gum (E415), guar gum (E412), carrageenan (E407), and pectin (E440) showing prevalence between 35–51%. PA additive class was led by ascorbic acid (E300) and potassium sorbate (E202) reaching 40–42%, while sodium nitrite (E250), sodium ascorbate (E301), and sulfur dioxide (E220) remained below 20%. SG showed moderate exposure in the 22–33% range, including acesulfame K (E950), sorbitol (E420), glycerol (E422), aspartame (E951), and sucralose (E955). Flavor enhancers were represented almost exclusively by monosodium glutamate (E621, 22%). Colorants, despite comprising the second highest number of unique additives in the study, showed low prevalence, with paprika extract (E160c), sulfite ammonia caramel (E150d), and carotenes (E160a) each reaching 17–23% of participants. These prevalences showcase that pH regulators and emulsifiers dominate the general additive exposure, reflecting current food industry practices focused on shelf-life extension, flavor control, and textural stability across different commercially available foods (Supplementary Figure 1a).

**Figure 1.**
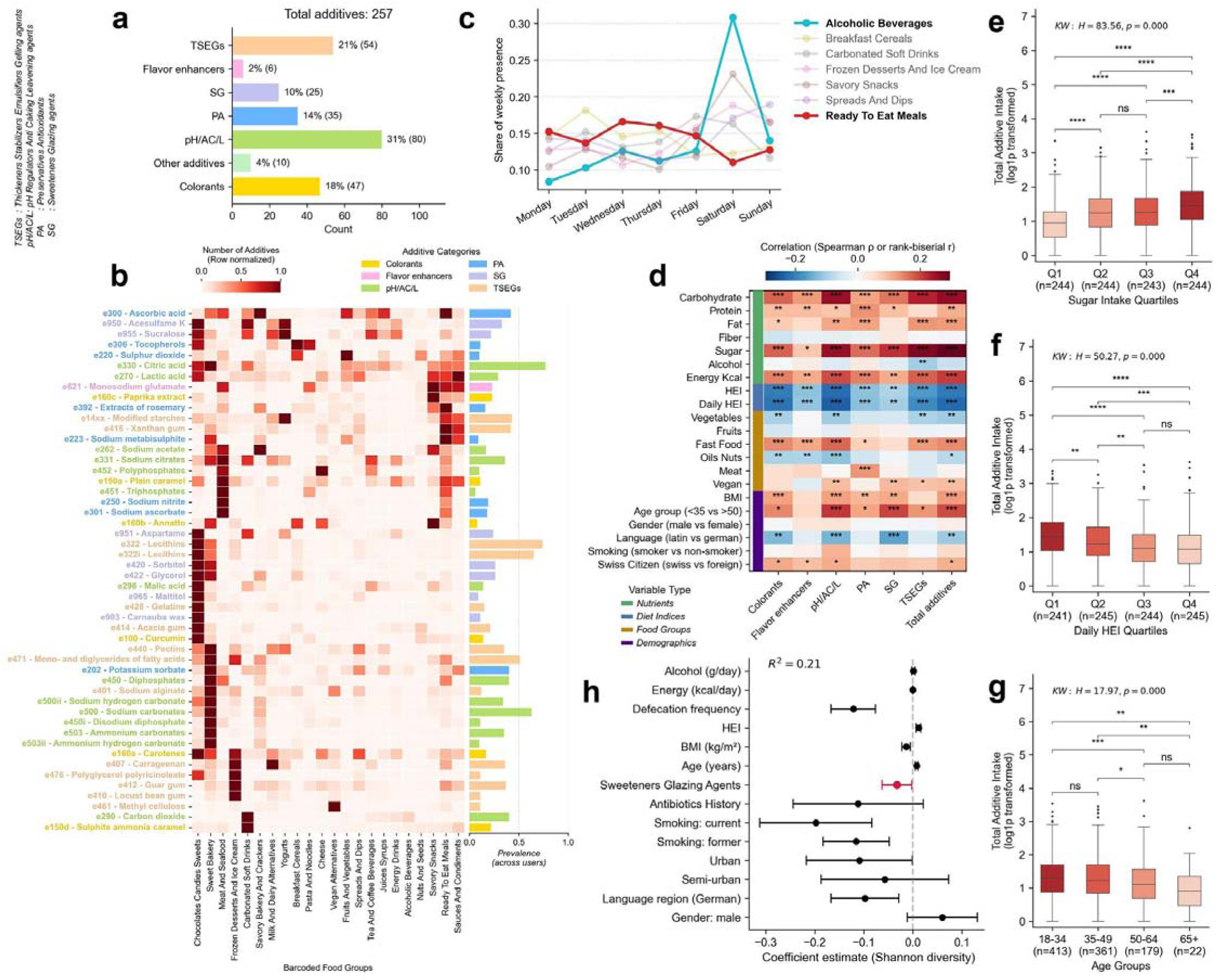
Food additive exposure across food groups, diet quality, and demographic factors. **(a)** Bar chart showing the proportion of unique additives by each additive category identified in the Food & You study. **(b)** Heatmap clustering of the top 50 additives (by frequency), showing the counts of additives (row normalized) within barcoded food groups. Bar plots on the right indicate additive prevalence across users, with colors denoting additive categories. Each additive is designated by an E-code, a standardized numerical identifier assigned under European food safety regulations. **(c)** Weekly distribution of additives containing food groups consumption (as relative share of weekly occurrences) for each food group. Food groups with diverging weekday versus weekend trends (alcoholic beverages and ready to eat meals) are highlighted. **(d)** Heatmap showing Spearman correlation coefficients for continuous predictors (dietary features) and rank biserial correlation coefficients from two-sided Mann–Whitney U tests for categorical predictors (age group, gender, language, smoking, citizenship), where positive values indicate higher additive exposure in the first listed category. Additive intakes were log1p transformed prior to testing. Positive values indicate higher additive exposure in the high quartile or first listed category. Significance was assessed using Benjamini–Hochberg false discovery rate correction across all tests, with asterisks indicating FDR-adjusted q-values (*q <; 0.05, **q <; 0.01, ***q <; 0.001). **(e–g)** Boxplots show total additive intake (log1p transformed) across quartiles of (e) sugar intake, (f) daily Healthy Eating Index, and (g) age groups. Group differences were assessed using Kruskal-Wallis (KW) non-parametric test. Sample sizes for each group are indicated in parentheses on the x-axis. Significance levels (ns; *p <; 0.05; **p <; 0.01; ***p <; 0.001; ****p <; 0.0001) are annotated on each panel. **(h)** Multivariable regression coefficients for the association between sweeteners-glazing agents intake and gut Shannon diversity (n = 940). Points represent coefficient estimates and horizontal bars indicate 95% confidence intervals. The additive term is highlighted in red.

Figure 1b displays the distribution of the 50 most frequently occurring additives across food categories. Additive occurrence was quantified per unique product and row-normalized to enable comparison across food categories with different sample sizes. Food categories with similar processing requirements (e.g., shelf-stable products and baked goods) clustered together and shared similar additive profiles. Sweet bakery products, chocolates/candies/sweets, and ready-to-eat meals exhibited the highest diversity of additives (exemplified for sweet bakery food group in Supplementary Figure 1b). pH regulators like citric acid and lactic acid were consistently present across most food categories, with particularly high representation in sweet bakery products, as well as in beverages and sauces and condiments. Meat and seafood products and ready-to-eat meals shared common preservatives including sodium nitrite (E250), sodium ascorbate (E301), and triphosphates (E451), while meat products additionally showed intensive usage of potassium nitrate (E252), polyphosphates (E452), and sodium citrates (E331). Ready-to-eat meals were further characterized by rosemary extracts (E392), sodium metabisulphite (E223), modified starches (E14xx), xanthan gum (E415), sodium acetate (E262), and monosodium glutamate (E621). TSEGs (e.g., xanthan gum (E415), mono- and diglycerides of fatty acids (E471)) showed high presence in dairy products, sweet bakery products, and frozen desserts, with vegan products showing high representation of methyl cellulose (E461). Colorants (carotenes (E160a), annatto (E160b)) were concentrated in sweet products, savory snacks, breakfast cereals, and cheese, while sweeteners such as sucralose (E955) and acesulfame K (E950) were primarily found in carbonated soft drinks, dairy/yogurts, and sweet products.

We also examined the temporal distribution of barcoded food group consumption across the week (Figure 1c and Supplementary Figure 2), as this provides insight into day-level variation in additive intake. We observed a weekend effect, with discretionary food groups such as alcoholic beverages, savory snacks, frozen desserts, juices and syrups, and spreads and dips all increasing toward the weekend; alcoholic beverages showed the strongest weekend-oriented pattern (Figure 1c). In contrast, breakfast cereals, yogurts, and milk and dairy alternatives displayed higher weekday consumption. Sweet bakery products, sauces and condiments, savory bakery and crackers, and tea and coffee beverages showed minimal day-to-day variation (Supplementary Figure 2).

Increased Consumption of Additives Associated with Lower Dietary Quality

To assess how intake of different food additive classes varies across dietary, lifestyle, and demographic characteristics, we computed Spearman correlations for continuous predictors and rank biserial correlations from Mann–Whitney U tests for categorical predictors (Figure 1d). Across nearly all additive classes, higher overall dietary intake and lower dietary quality were consistently associated with higher additive consumption.

Participants with higher intakes of carbohydrates, sugars, fats, and total energy showed significantly higher intakes across most additive classes. Sugar intake showed the strongest associations with total additive exposure (Kruskal-Wallis (KW): H = 83.56, p < 0.001; Figure 1e). Conversely, higher dietary quality, as measured by Healthy Eating Index (HEI) and daily HEI, was consistently associated with lower intake across nearly all additive classes (KW: H = 50.27, p < 0.001 for total additive exposure; Figure 1f). Higher fast food consumption was also associated with significantly higher intake across multiple additive classes, whereas vegetables intake showed only limited inverse associations while no differences were observed for fruits and fiber intakes.

With respect to additive classes, pH/AC/L exhibited the broadest and most consistent set of associations across dietary quantity, dietary quality, fast food intake, and demographic contrasts, indicating that this additive class may serve as a general marker of industrial food processing intensity. PA showed a strong association with meat consumption along with protein and fats, consistent with preserved animal based foods as a major contributor to this additive class. SG were characterized by very strong associations with sugar intake, but not with fast foods and meat food groups. TSEGs showed strong positive associations with sugar intake, fat intake, and fast food consumption, modest increase among vegan foods while modest negative relation with vegetables.

Differences in demographic and lifestyle factors were also prominent. Higher BMI and younger age were associated with higher intake of several additive classes, particularly SG, PA, colorants and pH/AC/L, with total additive intake also differing significantly across age groups (KW: H = 17.97, p < 0.001; Figure 1g). Participants from French and Italian speaking regions of Switzerland showed consistently lower additive exposure across several classes compared to those from German speaking regions (Supplementary Figure 3a). No significant differences in additive intake were observed by gender or smoking status.

### Three Additive Mixtures Linked to Diet Quality and Demographics

While individual additive prevalence provides valuable insights, additives rarely occur in isolation within processed foods. Moreover, additives may perform multiple functions, complicating their assignment to specific predefined classes. For example, sodium carbonates act as both pH regulators and texture modifiers, while sorbitol serves as a sweetener and also functions as a humectant and bulking agent. We therefore employed non-negative matrix factorization (NMF) to identify naturally occurring patterns of additive co-consumption, which may better reflect real-world dietary exposures than examining additives individually. This approach has recently been used in a large-scale study to identify cardiometabolic disorder related causal patterns^15^. Therefore, this approach would help us to characterize functional additive mixtures that potentially have combined effects on gut microbiota composition beyond those of single additives. Based on cophenetic coefficient analysis across multiple initializations, we identified three distinct patterns (i.e., mixtures) of additive co-occurrence (also shown in Figure 2a), and these mixtures also align with different processed food categories (Figure 2b).

**Figure 2:**
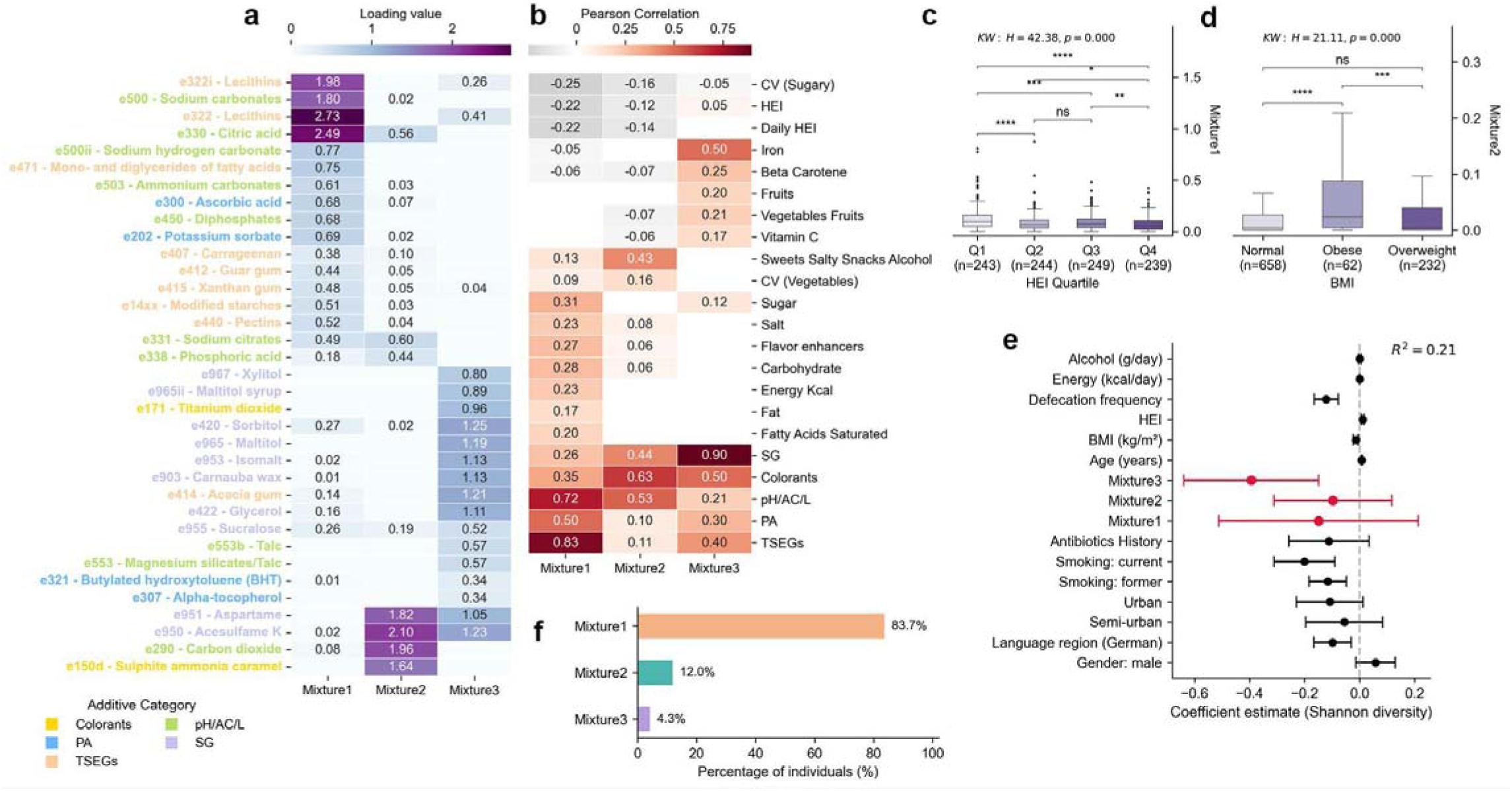
Additive mixture patterns and their associations with diet, demographics, and gut microbiota diversity. **(a)** Heatmap illustrating the distribution and contribution of E-coded additives across three distinct mixture components identified via NMF. Color intensity represents loading magnitude, with darker purple indicating stronger contributions. Only additives with loadings >0.3 in at least one mixture are displayed. These mixtures represent co-occurring additive patterns in ultra-processed foods: Mixture 1 (emulsifiers + pH regulators), Mixture 2 (sweeteners + colorants + pH regulators), and Mixture 3 (sweeteners + glazing agents + sugar polyols). Numerical values indicate the specific loading of each additive within its respective mixture**. (b)** Heatmap displaying Pearson correlation coefficients between the three identified additive mixtures and various nutritional and dietary components. Only correlations with absolute values ≥ 0.1 are displayed. The color intensity represents correlation strength, with darker red indicating stronger positive correlations and gray indicating negative correlations. Panels **(c–d)**: Boxplot showing relationship between mixture1 across HEI quartiles (c) and mixture2 across BMI categories (d). Statistical tests (KW = Kruskal–Wallis; MW = Mann–Whitney U) and significance levels (ns; *p <; 0.05; **p <; 0.01; ***p <; 0.001; ****p <; 0.0001) are annotated on each panel. **(e)** Multivariable regression coefficients for the association between additive mixtures and gut Shannon diversity (n = 940). Points represent coefficient estimates and horizontal bars indicate 95% confidence intervals. Mixture terms are highlighted in red; the dashed vertical line denotes a null effect. **(f)** Distribution of dominant additive mixture types across the study population. The chart displays the percentage of participants for whom each mixture represents the highest component score.

Figure 2a displays the loading values of emblematic additives across the three identified mixtures, wherein each mixture represents functional groupings based on additive contributions (Supplementary Table 2). Mixture1 was predominantly characterized by emulsifiers and acidity regulators, with the highest loadings observed for Lecithins, Citric acid, and Sodium carbonates, all with loadings ≥1.8. Mixture2 was defined by sweeteners, carbonation, colorants, and pH regulators, notably Acesulfame K, Carbon dioxide, Aspartame, and Sulphite ammonia caramel, with loadings ranging from 1.6 to 2.1, indicating a pattern consistent with flavored and carbonated beverage consumption. Mixture3 comprised mainly sweeteners-glazing agents, including Acesulfame K, Aspartame, and Carnauba wax (E903), as well as sugar alcohols such as Sorbitol, Maltitol (E965), and Isomalt (E953), all with loadings >1.0. Some pH regulators like Citric acid, Sodium citrate and Phosphoric Acid (E338) showed cross-contributions between mixture1 and mixture2.

To understand the nutritional context of the identified additive mixtures, we analyzed their Pearson correlations with dietary factors and nutrient intakes (Figure 2b). Mixture1 showed strong positive correlations with TSEGs and pH/AC/L (r=0.72–0.83), alongside moderate associations with sugar, carbohydrates, and flavor enhancers (r=0.27–0.32), consistent with processed carbohydrate-rich foods. Mixture2 demonstrated strong correlations with colorants and pH/AC/L (r=0.53–0.63). Mixture3 showed the strongest correlation with SG (r=0.90), followed by colorants, TSEGs, and iron (r=0.40–0.50). Both mixtures 1 and 2 showed negative correlations with irregular consumption of sugary foods (CV sugary i.e., coefficient of variation of daily sugary food intake) and with HEI (KW: H = 42.38, p < 0.001; Figure 2c), suggesting these patterns may be associated with lower overall diet quality. Mixture2 exposure was also higher among participants with elevated BMI (KW: H = 21.11, p < 0.001; Figure 2d) and among German-speaking participants compared to French- or Italian-speaking regions (MW: U = 91,506, p < 0.001). Interestingly, mixture3 showed positive correlations (Figure 2b) with beta carotene, vitamin C, vegetable-fruit intake (r=0.17–0.25), and iron (r=0.50), but also had marginally higher exposure among physically active participants (>3 times/week; KW: H = 9.26, p = 0.010). This co-occurrence of sweetener-glazing agent consumption with otherwise health-conscious dietary and lifestyle patterns may reflect deliberate use of artificially sweetened or “light” products as lower-calorie alternatives among individuals actively managing their diet and weight.

### Sugar Polyol- and Sweetener-rich Additive Mixture Associated with Reduced Gut Microbial Diversity

Given that additive exposures were associated with both dietary and demographic factors, we constructed multivariable linear regression models to assess whether associations with gut microbiota diversity were independent of these potential confounders. Models were adjusted for age, BMI, gender, HEI, language region, defecation frequency per day, urbanity, antibiotic treatment in the past six months, alcohol consumption, smoking status, energy consumed (daily average), and batch effects (n = 940).

In models examining individual additive classes using OLS with heteroskedasticity-consistent (HC3) standard errors, both sweeteners-glazing agents (β = −0.033, p = 0.035; Figure 1h) and preservatives-antioxidants (β = −0.085, p = 0.007; Supplementary Figure 3b) were negatively associated with Shannon diversity. Concordantly, mixture3 showed a significant negative association with Shannon diversity in standard OLS models (β = −0.39, 95% CI: −0.64 to −0.15, p = 0.002), however, mixtures 1 and 2 were not significantly associated (Figure 2e). This effect strengthened in a robust regression model using Huber loss (β = −0.48, 95% CI: −0.73 to −0.24, p < 0.001) and remained significant after adjustment for diet quality, BMI, and lifestyle covariates, indicating that it is independent of healthier dietary patterns that co-occur with additive exposure.

In the mixture model, age (β = 0.0077, p < 0.001) and HEI (β = 0.011, p < 0.001) were positively associated with microbial diversity, whereas BMI (β = −0.014, p = 0.002), defecation frequency per day (β = −0.12, p < 0.001), and German-speaking region (β = −0.10, p = 0.005) were negatively associated (Figure 2e). Similar effect sizes were seen in other additive models. Overall, these models explained approximately 20% of the variance in Shannon diversity.

Gut microbiota beta diversity was significantly associated with additive intake across multiple categories (PERMANOVA on unweighted UniFrac distances, marginal models adjusted for the core covariate set described above). Total additive intake showed the strongest association (R² = 0.0024, p = 0.002), followed by SG (R² = 0.0021, p = 0.005), PA (R² = 0.0019, p = 0.005), TSEGs (R² = 0.0017, p = 0.013), and pH/AC/L (R² = 0.0015, p = 0.027). No evidence of differences in within-group dispersion was observed for any additive category (PERMDISP all p > 0.5). Consistent to above observations, additive classes and mixtures aligned along the same primary compositional gradient, whereas fruit and vegetable intake vectors pointed in the opposite direction, (Supplementary Figure 3c).

### Vegetable benefits for microbiome diversity are counteracted by high additive intake

Given the independent negative associations between additive exposure and gut microbial diversity, we next examined whether additive intake modifies the relationship between protective dietary components (vegetable and fruit intake) and microbial diversity. We fitted multivariable linear regression models with Shannon diversity as the outcome (Figures 3a-b), adjusting for the core covariate set described in Methods, with the exception of HEI, which was excluded to avoid collinearity with the dietary variables being tested. All dietary variables were standardized (z-scored) to facilitate comparison of effect sizes, and robust (HC3) standard errors were used throughout.

**Figure 3:**
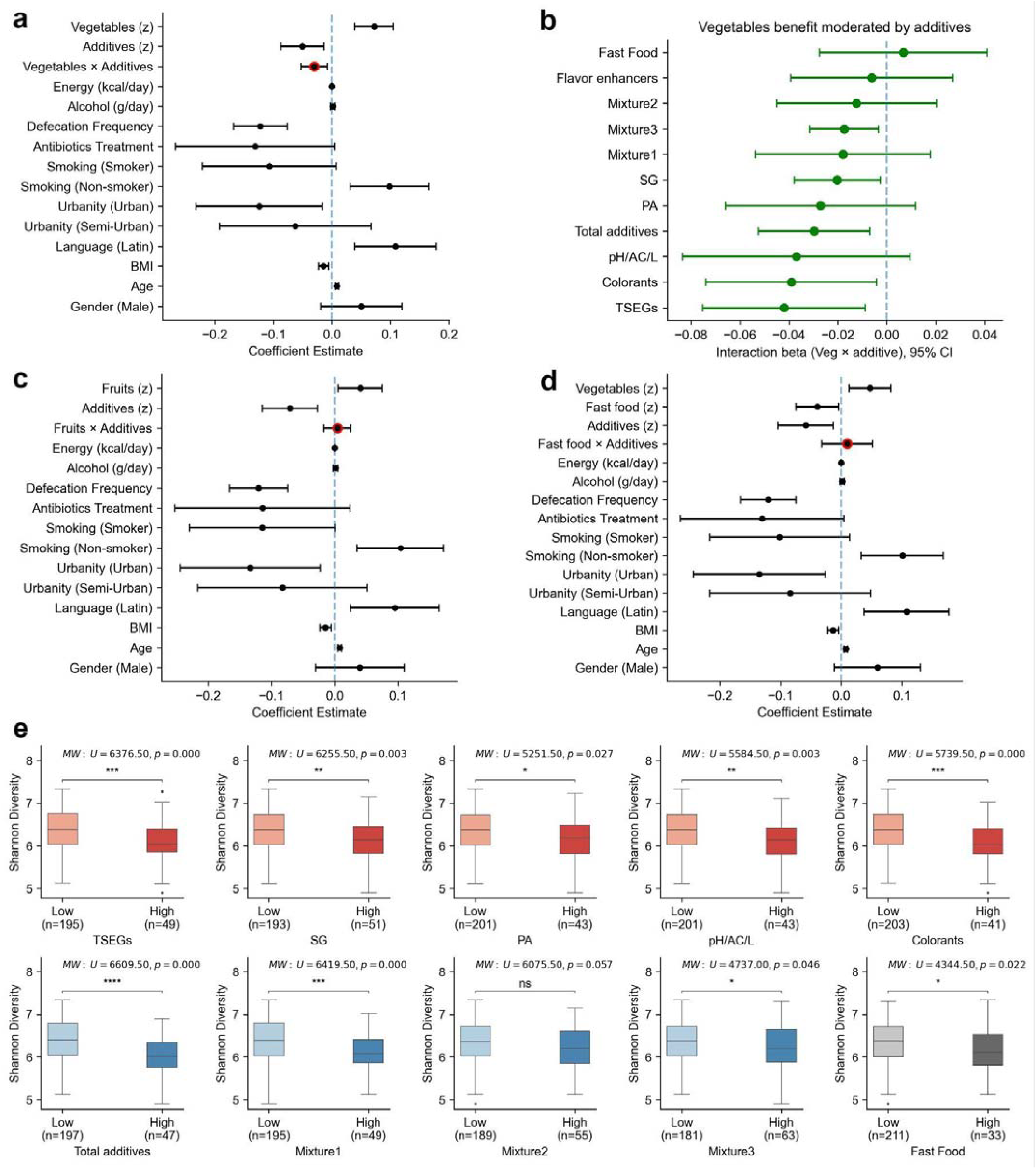
Food additive intake modifies the relationship between vegetable consumption and gut microbiome diversity. **(a)** Forest plots of multivariable linear regression coefficients predicting gut microbiota Shannon diversity for the vegetable model (n = 940). The model includes: demographics (age, BMI, gender), geographic factors (language, urbanity), lifestyle behaviors (smoking, antibiotics treatment), defecation frequency per day and dietary factors. Vegetables and total additive intakes are z-standardized; coefficients for these variables represent the change in Shannon diversity per one standard deviation increase. Non-standardized variables display units in brackets. The vegetable × total additive interaction terms are highlighted in red. Error bars represent 95% confidence intervals. **(b)** Forest plots showing vegetable × additive interaction coefficients. Each point represents the interaction term from separate multivariable regression models testing interactions between vegetable intake and different additives and fast food, adjusted for covariates. Negative coefficients indicate that additive intake attenuates the association between vegetables and increased gut alpha diversity. Error bars: 95% confidence intervals. **(c)** Forest plot for the fruit model, structured as in panel a, with the fruit × total additive interaction term highlighted in red. **(d)** Forest plot for examining the interaction between fast-food consumption and total additive intake. Includes same covariates as panels a and c, but also includes scaled vegetable intake. **(e)** Boxplots compare Shannon diversity between high vegetable consumers (>75^th^ percentile) with low versus high intake of various additives, mixtures and fast food consumption. Top row (red) shows different additive categories, while the bottom row shows total additive intake, additive mixtures and fast food intake. Mann-Whitney U test statistics and p-values are displayed above each plot, with significance indicated (*p <; 0.05, **p <; 0.01, ***p <; 0.001, ****p <; 0.0001, ns=not significant). Sample sizes for each group are shown in parentheses.

In the vegetable model (Figure 3a), vegetable intake was positively associated with microbial diversity (β = 0.069, 95% CI: 0.036 to 0.101, p < 0.001), while total additive intake showed an independent negative association (β = -0.052, 95% CI: -0.089 to -0.015, p = 0.005). Importantly, a significant negative vegetable by additive interaction was observed (β = -0.030, 95% CI: -0.052 to -0.007, p = 0.010), indicating that the positive association between vegetable intake and microbial diversity weakened with increasing additive exposure. To identify which additive categories most strongly drive this interaction, we examined interaction terms between vegetable intake and individual additive categories (Figure 3b). Several additive categories showed significant negative interactions with vegetable intake, indicating that higher exposure to specific additives consistently weakened the diversity-associated benefit of vegetables. The strongest interaction was observed for TSEGs (β = -0.042, p = 0.013), followed by colorants (β = -0.039, p = 0.028), SG (β = -0.020, p = 0.024), and mixture3 (β = - 0.018, p = 0.014). Other categories including preservatives and antioxidants, pH/AC/L, flavor enhancers, and mixtures, showed consistent negative interaction estimates but did not reach statistical significance.

In contrast, in the fruit model (Figure 3c), fruit intake was independently and positively associated with Shannon diversity (β = 0.038, 95% CI: 0.004 to 0.073, p = 0.030), while total additive intake remained negatively associated (β = -0.072, 95% CI: -0.115 to -0.028, p = 0.001). However, no evidence of interaction between fruit intake and additive exposure was observed. Effects of covariates were consistent across both models and aligned with previous analyses (Figures 1h and 2d).

Given that fast food consumption often co-occurs with high additive exposure in Western dietary patterns, we next tested whether fast food intake exhibits a similar interactive effect with additive exposure on gut microbial diversity. We fitted a multivariable linear model including a fast food by total additive interaction, while additionally adjusting for vegetable intake (z-scored) to isolate effects independent of protective food consumption. In this model, both dietary factors showed independent negative associations with Shannon diversity (Figure 3d): fast food intake was associated with lower diversity (β = -0.034, 95% CI: -0.068 to 0.000, p = 0.052), while additive intake showed a strong negative association (β = -0.060, 95% CI: -0.106 to -0.015, p = 0.010). However, no evidence of interaction between fast food and additive intake was observed (β = 0.010, 95% CI: -0.033 to 0.052, p = 0.661), indicating that fast food and additives are associated with reduced microbial diversity through additive, but not synergistic, effects. This pattern contrasts with the vegetable by additive interaction, where higher additive exposure specifically attenuated the microbiome benefits associated with vegetable intake.

To isolate the impact of additives among individuals already consuming protective foods, we stratified participants with high vegetable intake (top quartile) by additive exposure levels (high vs low, defined by the 75^th^ percentile of each additive). This approach allowed us to directly test whether high additive exposure attenuates microbiome benefits even within a vegetable-rich dietary context. Within this high-vegetable subgroup (Figure 3e), participants with high additive exposure consistently exhibited lower gut microbial alpha diversity compared to their low-additive counterparts. The strongest associations were observed for total additive intake (p < 0.001), TSEGs (p < 0.001), colorants (p < 0.001), and SG (p = 0.003). Additional significant reductions in diversity were seen for pH/AC/L (p = 0.003) and PA (p = 0.027). Among additive mixtures, mixture1 showed a significant association with reduced diversity (p < 0.001), mixture3 showed a weaker but significant effect (p = 0.046), while mixture2 was not significant. High fast food consumption was also associated with reduced microbial diversity within this high-vegetable group (p = 0.022). Applying the same stratification to high fruit consumers revealed fewer and weaker associations (for PA and pH/AC/L; Supplementary Figure 3d). This indicates that the microbiome benefits associated with vegetable intake may be more vulnerable to disruption by concurrent additive exposure than those associated with fruit intake.

### Microbiota Log Ratios of Additives Show Strong Associations with Gut Diversity

To identify gut microbes that increase or decrease in abundance with additive exposure, we constructed microbial log-ratio metrics that summarize community-level responses. These log ratios were calculated by dividing the summed abundances of taxa positively associated with each feature of interest (e.g, an additive) by those negatively associated, determined via differential abundance analysis using BIRDMAn^20^. Resulting log ratios reflect the balance of microbial responders linked to a specific additive, or dietary feature like HEI.

By correlating these microbial signatures with alpha diversity, we can assess whether the specific taxa that respond to additive exposure are themselves linked to community-level diversity, thereby connecting additive-associated compositional shifts to overall ecosystem structure. Figure 4a shows a correlation heatmap between microbial log ratios and various alpha diversity metrics, highlighting a consistent pattern of negative associations between additive associated log ratios and gut diversity. These correlations ranged from approximately r = -0.35 to -0.55 for total additive intake across gut alpha diversities. Among individual additive groups, TSEGs exhibited the strongest inverse relationship with gut diversity, including Shannon diversity (≈r = -0.65), with similarly strong negative associations observed for fast food, colorants, SG and pH/AC/L. These log ratios were similarly negative to HEI score (Figure 4b), most notably for pH/AC/L additives (r = -0.35, p = 2.56 × 10⁻²⁹) and PA (r = -0.20, p = 5.25 × 10⁻¹⁰). Log ratios for additive mixtures also showed negative associations with gut diversity, wherein mixture3 exhibited the strongest negative association with Shannon diversity (r = -0.49).

**Figure 4:**
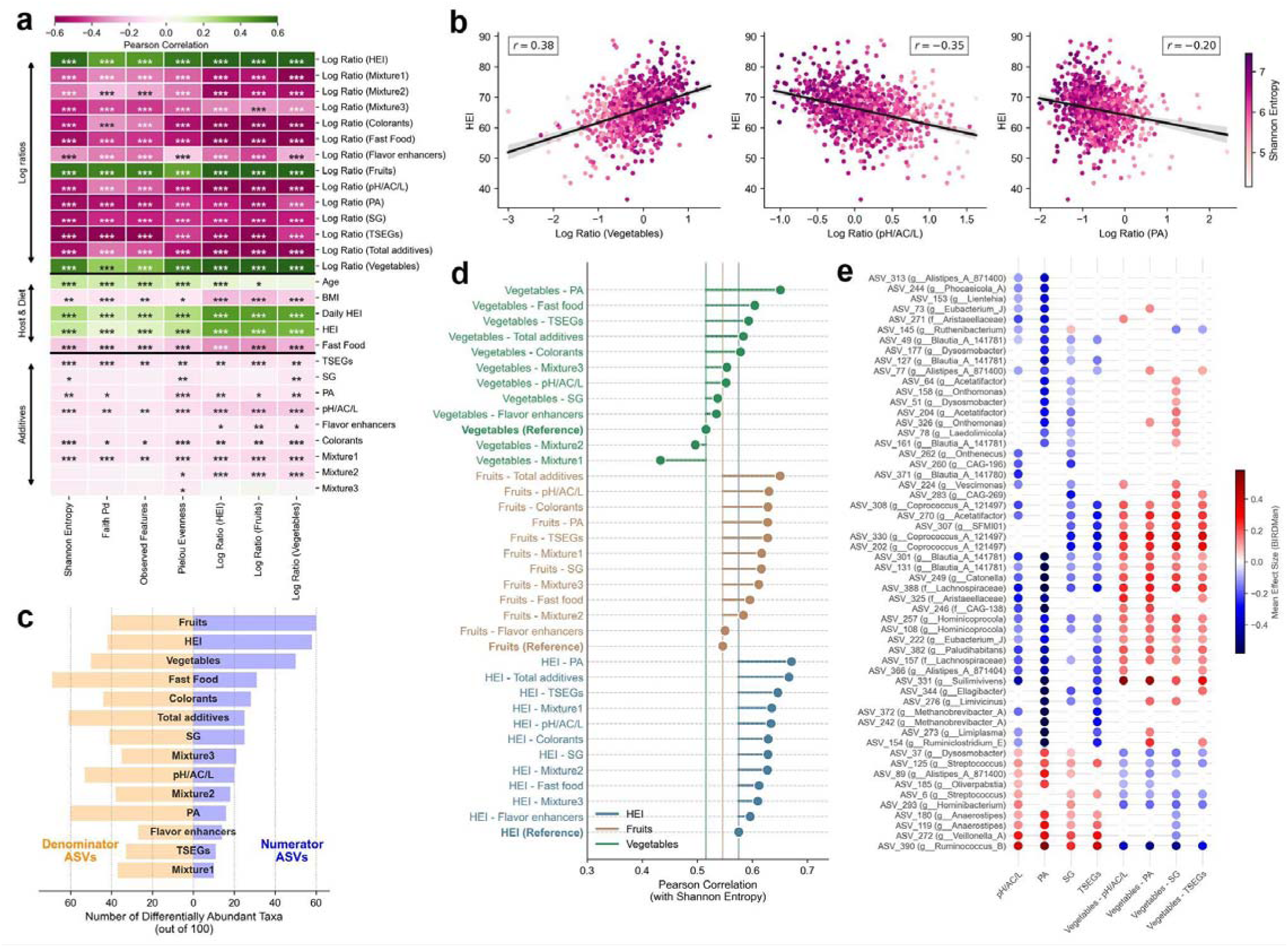
Microbial log-ratio metrics reveal stronger correlations between dietary quality and additive exposure. **(a)** Pearson correlation heatmap showing relationships between food additive exposures, microbial log ratios, host factors, and gut microbiota alpha diversity metrics. Log ratios represent the relative abundance of microbes positively versus negatively associated with each additive or food group. Color intensity indicates correlation strength. Significance levels are denoted by asterisks (*p <; 0.05, **p <; 0.01, ***p <; 0.001). **(b)** Scatter plots showing negative correlations between HEI scores and microbial log ratios associated with intakes of additives and vegetables. Points are colored by Shannon diversity (lighter to darker purple representing increasing diversity). Pearson correlation (r) is highlighted in each panel. Gray shading represents 95% confidence intervals for regression lines. **(c)** Mirror barplot showing the number of differentially abundant taxa associated with dietary factors and food additive exposures. Blue bars (right) indicate taxa enriched in the numerator (positively associated), while orange bars (left) indicate taxa enriched in the denominator (negatively associated) for each log-ratio metric. **(d)** Correlations between Shannon diversity and microbial log ratios derived from composite dietary metrics. Composite metrics were defined as the z-score difference between a healthy dietary component (HEI, fruits, or vegetables) and an additive or fast food variable (e.g., HEI-minus-PA). Microbial log ratios were then constructed from BIRDMAn differential abundance analysis on these composite metrics. Reference lines indicate base correlations for log ratios of dietary components alone. Correlations are color-coded by reference group (blue = HEI, brown = fruits, green = vegetables). **(e)** Heatmap displaying mean effect sizes from BIRDMan differential abundance analysis for taxa significantly associated with food additive and corresponding composite metrics. Color indicates the effect direction and magnitude (blue = negative association, red = positive association). Only taxa with significant associations across multiple variables are shown.

In contrast, positive correlations were observed between alpha diversities and the log ratios of fruit and vegetable consumption (r = 0.30 to 0.55), and with HEI log ratio showed (r = 0.45 to 0.60), reinforcing the link between overall diet quality and microbiome health^21,22^.

Examining the composition of each log ratio in terms of contributing taxa (Figure 4c), we see that HEI, fruits, and vegetables were derived from a larger set of differentially abundant taxa and exhibited balanced or numerator-enriched profiles. Additive-related features and fast food consumption, in contrast, were characterized by fewer responding taxa overall and a consistent dominance of negatively associated taxa, indicating a net depletion of microbial diversity in relation to additive-rich dietary exposures.

### Enhanced Microbiota Associations of Combined Dietary Quality and Additive Metrics

To investigate the balancing effects between health-promoting dietary components versus harmful ones (i.e., additive or fast foods) on gut diversity, we developed contrasting metrics using standardized scores. For each healthy dietary variable (HEI, fruits, and vegetables), we calculated the difference between its z-score and that of each additive, creating “minus” variables that quantify the relative dominance of beneficial over potentially detrimental ones. We then constructed microbial log ratios for these composite metrics, following the same approach used for individual dietary features. If additive exposure attenuates the microbial benefits of healthy diets, these composite log ratios should highlight stronger associations with gut diversity than either component alone.

As shown in Figure 4d, HEI-minus-PA and HEI-minus-total additive intake showed the strongest correlations with Shannon diversity (both r ≈ 0.67), exceeding the baseline HEI correlation (r = 0.58). Similar gains were observed for fruit-based composite metrics, particularly fruits-minus-total additive intake and fruits-minus-PA (r ≈ 0.63–0.65), indicating that accounting for additive burden substantially strengthened associations with gut diversity. While vegetables-minus-PA and vegetables-minus-fast food improved relative to the baseline vegetable association, other vegetable-minus metrics showed weaker improvements, and for some, such as mixture1 and mixture2, correlations were actually reduced compared to reference.

### Taxonomic comparison across differential additive signatures

We compared the mean effect sizes of differentially abundant taxa across additive and vegetables-minus contrasts (Figure 4e). From taxa significantly associated with at least one of three additive classes (SG, TSEG, or pH/AC/L), we retained those with summed absolute effect sizes > 0.25 across additive features.

Commensal and fiber-associated taxa showed negative effect sizes under additive exposure but positive effect sizes under vegetables-minus features, indicating a consistent directional shift depending on dietary context. These included Lachnospiraceae (ASV_388, ASV_157), Coprococcus_A_121497 (ASV_330, ASV_308, and ASV_202), Eubacterium_J (ASV_222), and Blautia_A_141781 (ASV_301 and ASV_131). Across these taxa, vegetables-minus effect sizes showed a median Δ ≈ +0.25 (IQR ≈ 0.16 to 0.28), whereas additive-only effects were negative, with a median Δ ≈ -0.16 (IQR ≈ -0.29 to -0.16). This directional shift is consistent with enrichment of these taxa in vegetable-rich dietary contexts relative to additive burden. Notably, PAs showed some of the strongest additive-only depletions, including Ruminiclostridium_E (ASV_154, Δ = -0.90), Methanobrevibacter_A (ASV_372, Δ = -0.71; ASV_242, Δ = -0.62), Catonella (ASV_249, Δ = -0.64), and CAG-138 (ASV_246, Δ = -0.63), with corresponding positive shifts under vegetables-minus-PA (e.g, ASV_249, Δ = +0.26; ASV_154, Δ = +0.24).

In contrast, some facultative or opportunistic taxa displayed the opposite pattern, characterized by enrichment under additive-only conditions and depletion under vegetables-minus contrasts, with PA often driving the strongest effects. Most notably, Ruminococcus_B (ASV_390) showed strong PA-linked enrichment (Δ = +1.12) alongside pronounced negative vegetables-minus effects (Δ = -0.49). Similar patterns were observed for Anaerostipes (ASV_119, PA Δ = +0.31), Veillonella_A (ASV_272, PA Δ = +0.35), and Alistipes_A_871400 (ASV_89, PA Δ = +0.28). Streptococcus (ASV_125) showed smaller but consistent PA-linked enrichment (Δ = +0.20) with negative shifts across vegetables-minus features (e.g., vegetables-minus-SG Δ = -0.14).

Collectively, these patterns indicate that additive exposure, for instance preservatives-antioxidants, is associated with enrichment of facultative or opportunistic taxa, while depleting core commensals linked to dietary fiber and metabolic health.

### Sweetener-associated enrichment of polyol and aromatic compound metabolism

To investigate potential functional mechanisms underlying the observed microbiome associations, we examined microbial metabolic pathways inferred using PICRUSt2^23^. Pathway relative abundances were tested for association with individual food additive groups and dietary mixtures using multivariable linear models implemented in MaAsLin2 and OLS models, adjusting for demographic, lifestyle, and technical covariates (see methods for details). For each exposure, models were fitted independently, and statistical significance was assessed using Benjamini–Hochberg correction across all tested pathway–exposure pairs.

Pathway associations were predominantly driven by sweeteners-glazing agents, which showed the highest count of significant pathways (46 pathways, derived from MaAsLin2), as shown in Figure 5a. Mixture3 showed 55 significant pathways, 43 of these were shared with SG and 12 pathways were unique to mixture3 (Figure 5b). In contrast, other individual additive categories exhibited substantially fewer associations, with some overlap with SG (11 shared pathways with colorants; 8 with preservatives and antioxidants).

**Figure 5.**
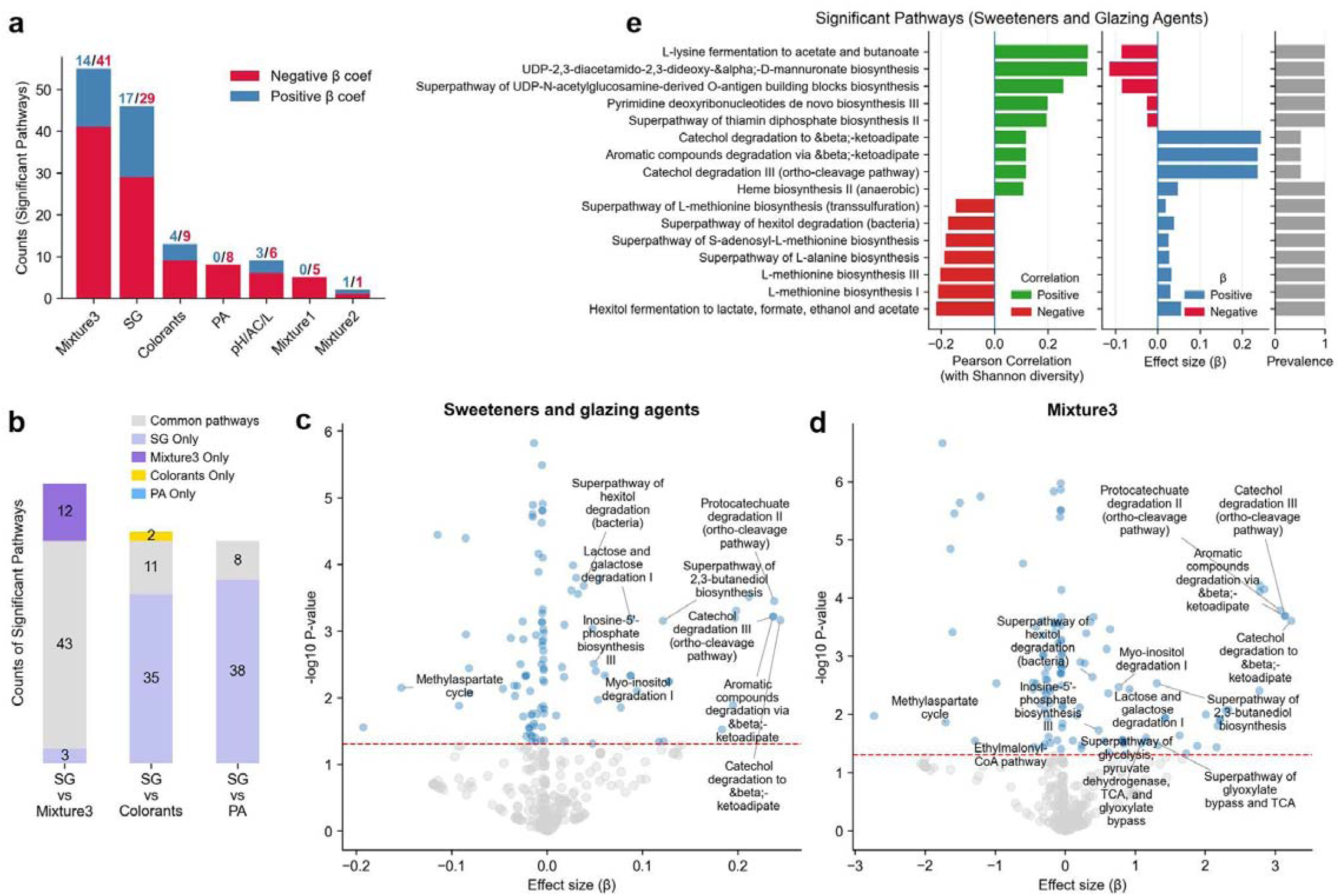
Predicted functional pathway associations with food additive exposures. **(a)** Counts of FDR significant PICRUSt2 predicted pathways associated with each additive exposure and mixture from MaAsLin2 models. Bars are stacked by coefficient direction (negative β in red, positive β in blue), and labels above bars indicate positive/negative counts for each exposure. **(b)** Overlap of FDR significant PICRUSt2 predicted pathways between SG and other exposures. Stacked bars show pathways unique to each exposure and pathways shared between each pair (gray segment). **(c-d)** Volcano plots of MaAsLin2 associations between PICRUSt2 predicted pathway abundances and (b) sweeteners and glazing agents and (c) mixture3. Points show pathway effect sizes (β) versus −log_10_(p value); the red dashed line denotes significance threshold of -log_10_(p = 0.05). Selected pathways are labeled based on overlap with OLS modeling results. **(e)** Shown are pathways that were FDR significant for sweeteners and glazing agents and exhibited an absolute Pearson correlation greater than 0.1 with Shannon diversity. Bars indicate correlation with Shannon diversity, corresponding MaAsLin2 effect sizes (β), and non-zero pathway prevalence across samples.

Both SG and mixture3 were positively associated with pathways involved in sugar polyol catabolism (Figure 5c-d, Supplementary Table 3), including the superpathway of hexitol degradation (HEXITOLDEGSUPER-PWY) and myo-inositol degradation I (P562-PWY). The strongest positive associations were observed for aromatic compound degradation pathways for β-ketoadipate (PWY-5431; PWY-5417; PROTOCATECHUATE-ORTHO-CLEAVAGE-PWY). SG and mixture3 were also linked to enrichment of fermentation and central carbon metabolism pathways, including 2,3-butanediol biosynthesis (PWY-6396), the glyoxylate cycle (GLYOXYLATE-BYPASS), TCA cycle IV (P105-PWY), and integrated superpathways linking glycolysis, TCA, and glyoxylate bypass (GLYCOLYSIS-TCA-GLYOX-BYPASS; TCA-GLYOX-BYPASS), which together support efficient carbon utilization during carbohydrate catabolism.

Conversely, SG exposure was negatively associated with the methylaspartate cycle (PWY-6728) and ethylmalonyl-CoA pathway (PWY-5741) (Supplementary Table 3). Both serve as alternative routes for acetyl-CoA assimilation to the glyoxylate cycle, and their depletion alongside glyoxylate cycle enrichment suggests a compositional shift favoring taxa utilizing the latter strategy. Ectoine biosynthesis (P101-PWY), which produces ectoine, an osmoprotectant and intestinal barrier stabilizer^24^, was negatively associated with SG exposure, possibly reflecting community shifts that is driven by the osmotic effects of dietary sugar polyols such as sorbitol.

Among the 46 MaAsLin2-derived significant pathways for SG, 16 showed notable correlations with Shannon diversity (|r| > 0.1). Pathways that were negatively associated with SG intake, such as L-lysine fermentation to acetate and butanoate, UDP-N-acetylglucosamine-derived O-antigen biosynthesis, pyrimidine deoxyribonucleotide de novo biosynthesis, and thiamin diphosphate biosynthesis, are positively correlated with diversity (r = 0.19–0.35, Figure 5e). Conversely, pathways positively associated with SG, such as catechol and aromatic compound degradation via beta-ketoadipate, hexitol degradation, and methionine/SAM biosynthesis, showed negative correlations with gut diversity (r = −0.12 to −0.22). This suggests that higher SG intake favors pathways linked to polyol and aromatic metabolism at the expense of core biosynthetic functions aligned with greater community diversity.

Together, these findings suggest that SG exposure is associated with the most pronounced shifts in microbial metabolic capacity among the additive categories examined.

### Predictive Performance Improves When Microbiota Models Integrate Processed and Healthy Food Signals

Given the observed microbiome-diet associations, we examined whether microbial profiles could accurately classify individuals based on their dietary patterns using machine learning models. We assessed model performance using both individual dietary features and composite metrics as prediction targets Specifically, we trained XGBoost classifiers on microbial species abundance data to discriminate between individuals with low (≤25^th^ percentile) versus high (≥75^th^ percentile) intake of each dietary variable. Model performance was quantified using the area under the receiver operating characteristic curve (AUROC), where values of 0.5 indicate no discriminatory ability. By comparing AUROC values across dietary feature targets, we assessed whether incorporating additive or processed food information enhanced the microbiome’s ability to reflect dietary patterns.

For vegetable consumption (Figure 6a), baseline classifiers using microbial profiles alone showed moderate performance (median AUROC = 0.76). Incorporating additive exposure, i.e., for composite metrics, the classification accuracy improved, wherein vegetables-minus-total additive intake achieved a median AUROC of 0.81, and vegetables-minus-fast food reaching higher performance (median AUROC = 0.84). A similar pattern was observed for fruit intake (Figure 6b), where baseline performance was lower (median AUROC = 0.74) but improved when accounting for additives and fast foods. Adjustment for total additive intake increased performance to a median AUROC of 0.78, while contrasting fruit intake against fast food consumption yielded the strongest discrimination (median AUROC = 0.80). In contrast, classification based on HEI diet quality already showed high baseline performance (Figure 6c), with only marginal improvements upon few contrasting features, namely, fast foods, pH/AC/L and colorants.

**Figure 6.**
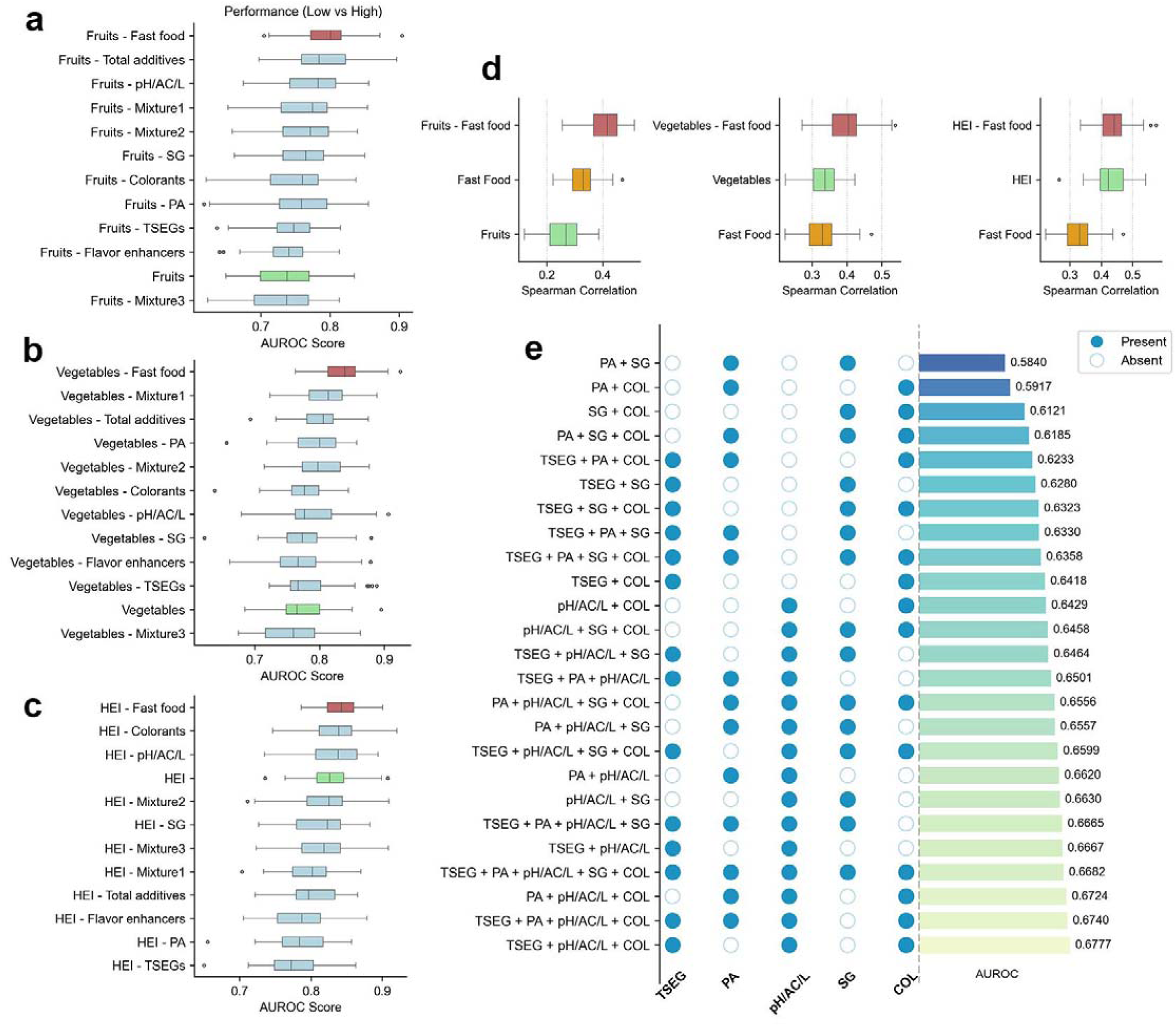
Machine learning prediction of dietary patterns from gut microbiota composition. **(a–c)** Area under the receiver operating characteristic curve (AUROC) scores from XGBoost classifiers trained on microbial species abundances to discriminate low (≤25^th^ percentile) versus high (≥75^th^ percentile) intake of vegetables (a), fruits (b), and HEI (c), as well as their composite metrics contrasting healthy dietary components against additive or fast food exposure. Baseline dietary variables are highlighted in green, fast food-adjusted metrics in red, and additive-adjusted metrics in blue. Higher AUROC values indicate improved ability of gut microbiota to distinguish dietary patterns. **(d)** Spearman correlation (ρ) between observed and predicted values from XGBoost regression models trained on microbial features to predict continuous dietary intake measures, with and without fast food adjustment. **(e)** AUROC performance for classifying high versus low exposure to different additive combinations. Presence/absence of each additive category is indicated by filled/empty circles (left), with overall classification performance shown by horizontal bars (right). Combinations are ranked by AUROC, with highest-performing combinations at the bottom.

Regression models fitted on the full cohort to predict continuous intake values corroborated these findings (Figure 6d). Incorporating fast food contrasts improved predictions for fruit intake (Spearman ρ increasing from 0.26 to 0.41) and vegetables (ρ = 0.33 to 0.40). For HEI, baseline performance was already high (ρ = 0.43), with minimal improvement upon adjustment (ρ = 0.44). These results suggest that microbiome composition reflects the net balance of dietary exposures rather than individual components in isolation.

Classification models for individual additive categories showed weak performance; we therefore performed a combinatorial analysis to test whether microbial profiles could discriminate between individuals in the lowest versus highest quartiles of summed additive exposure (Figure 6e). The highest performance was observed for combinations including TSEGs, pH/AC/L, and colorants (max mean AUROC = 0.68), with pH/AC/L appearing consistently across the top performing combinations, while adding PAs yielded comparable but not superior performance. In contrast, combinations involving PAs and TSEGs in the absence of pH/AC/L showed the weakest discrimination (mean AUROC ∼0.58). Classification based on total additive intake alone performed modestly (AUROC ∼0.65), even when restricting to extreme exposure quartiles. Overall, these results indicate that although certain additive combinations provide small gains in discrimination, microbiome based classification of additive exposure remains limited, particularly compared with the substantially stronger discrimination observed when contrasting healthy food intake with fast food or additive exposure, reinforcing that gut microbiome composition reflects integrated dietary patterns rather than additive burden in isolation.

## Discussion

Our study demonstrates that real-world exposure to mixtures of additives is associated with reduced gut microbial diversity in a free-living population. Unlike traditional FFQs that capture only broad food categories through retrospective recall, our high-resolution real-time dietary tracking in over 1,000 Swiss adults identified 257 specific additives from actual barcoded products consumed. This enabled us to discern dominant additive mixtures and found that their exposure patterns correlated with distinct demographic characteristics, dietary quality, and microbiota profiles. These findings build upon existing experimental and epidemiological literature by extending the relevance of additive-induced dysbiosis to population-level settings and highlighting the importance of considering additive mixtures, rather than single compounds, in microbiome research.

Mixture 1, which is dominated by emulsifiers (e.g., mono- and diglycerides, lecithins) and acidity regulators, was the most common exposure pattern in our cohort. The majority of participants (83.8%) had mixture1 as their dominant additive exposure pattern, i.e., additive mixture with the highest score (Figure 2f). This predominance is consistent with current food processing practices and aligns with experimental evidence showing that several commonly used emulsifiers and gums, including xanthan, guar, and carrageenans, induce lasting reductions in microbial diversity and pro-inflammatory shifts in ex vivo human microbiota models^25^. In our cohort, higher intake of TSEGs and pH/AC/L was associated with lower microbial diversity even among individuals with high vegetable intake, indicating that additive exposure remains detectable within otherwise plant-rich dietary contexts.

Moreover, beyond these independent associations, we observed clear evidence of effect modification, whereby additive exposure altered the relationship between plant-based food intake and gut microbiota diversity (Figure 3). While both vegetable and fruit intake were positively associated with Shannon diversity, higher additive consumption selectively attenuated the association for vegetables but not for fruits. This negative vegetable × additive interaction suggests that additive exposure may interfere with microbiota-accessible fibers or phytochemicals that otherwise support diversity-promoting taxa. In contrast, fast food and additive intake showed independent, non-synergistic associations with reduced diversity, implying partially distinct pathways of microbiome disruption. Together, these results indicate that the microbiome response to plant-rich diets depends not only on the quantity of vegetables consumed but also on the broader processed food context in which they are eaten.

Therefore, capturing the co-consumption of protective and detrimental dietary components is critical for characterizing the dietary gradients reflected in gut microbiome diversity. We addressed this by constructing composite “minus” metrics that explicitly contrast health-associated dietary components (e.g., HEI, fruit, and vegetable intake) against additive exposure. These contrasts, such as HEI-minus-total additive intake and vegetables-minus-preservatives, showed stronger associations with microbial diversity (r > 0.65) than dietary or additive measures considered in isolation. Consistently, microbiota-based classifiers and regressors showed improved performance when additive or processed-food information was incorporated alongside traditional dietary variables, highlighting the value of modeling opposing dietary exposures jointly rather than separately.

Notably, mixture3, characterized by high intensity sweeteners (acesulfame K, aspartame, sucralose) together with sugar polyols (e.g., sorbitol, maltitol, isomalt, xylitol), was negatively associated with Shannon diversity despite its low prevalence in the cohort (dominant mixture in ∼5% of participants; Figure 2f). Interestingly, mixture3 showed positive associations with micronutrients, fruit and vegetable intake, and physical activity, suggesting alignment with otherwise health-oriented dietary behaviors. This additive combination likely captures consumption of low-calorie, sugar-reduced or high protein products (e.g, beverages, supplements), a rapidly growing dietary segment that frequently relies on artificial sweeteners and sugar polyols to enhance palatability while maintaining low energy density. Although often marketed as healthier alternatives, such formulations may perturb the gut microbiome, either broadly or in a host-dependent manner.

Consumption of Splenda (sucralose/maltodextrin) has been shown to increase gut Proteobacteria and inflammation markers only in mice genetically prone to intestinal inflammation, suggesting that sweetener effects may be contingent on host susceptibility^26^. Sugar polyols are known to reach the colon due to incomplete absorption and can alter luminal osmotic load and fermentation dynamics in a dose-dependent fashion, with suggestive evidence of microbiome shifts in healthy individuals^13^. Although they can boost beneficial bacteria^14^, e.g., Bifidobacterium at moderate doses, higher exposures provoke laxation, gas, and broad compositional shifts^14,27^. Our findings suggest that exposure to different additive classes, including SG, in our cohort was associated with depletion of commensal and fiber associated taxa, including Lachnospiraceae, Coprococcus_A, Eubacterium_J, Blautia_A, Ruminiclostridium_E, and Methanobrevibacter_A, while pro-inflammatory taxa such as Ruminococcus_B and Veillonella_A were enriched.

Similarly, PICRUSt2-based functional profiling showed depletion of core biosynthetic and amino acid fermentation pathways with sweetener and glazing agent exposure, accompanied by enrichment of polyol catabolism and central carbon metabolism pathways. This pattern, together with reduced capacity for osmoprotectant biosynthesis (ectoine) and a shift from alternative acetyl-CoA assimilation routes toward the glyoxylate cycle, suggests that sugar alcohol-rich additive exposure may selectively favor taxa specialized in polyol degradation at the expense of functionally diverse commensal communities.

Long-term consumption of additive-rich dietary patterns may therefore have implications for metabolic health^15,28^, potentially mediated by changes in gut microbiota composition. In the NutriNet Santé cohort, comparable additive mixtures were associated with an increased risk of type 2 diabetes^15^, while in our study the same mixtures were linked to reduced gut microbial diversity. This comparison is supported by the strong similarity in additive exposure profiles across cohorts, including the high prevalence of additives such as citric acid and sodium carbonates, and the identification of emulsifier- and sweetener-dominated mixtures using NMF factorization in both populations^15,17^. Together, these converging observations point to a possible connection between additive exposure, microbiota disruption, and metabolic disease risk. Future longitudinal and intervention studies are needed to clarify the directionality and causality of these relationships.

While additives are commonly found in fast food, we observed only weak correlations between fast food intake and additive mixtures (r = 0.15 for mixture1, r = 0.07 for mixture2, r = -0.05 for mixture3). Although our fast food classification is based on food item type (e.g., pizza, hamburgers, salty snacks) rather than the source or preparation context, and therefore does not distinguish between commercially and home-prepared versions of the same item, these weak correlations indicate that the additive mixtures are not merely a proxy for fast food consumption patterns, and may affect gut microbiota mechanistically.

Furthermore, UPFs consumption is inherently tied to additive exposure, as NOVA 4 foods correlated moderately to strongly with several additive classes in our cohort (r = 0.28–0.60; Supplementary Figure 4a). However, in multivariable models simultaneously adjusting for all four NOVA processing categories and the full covariate set, total additive intake (□ = −0.065, p = 0.024), preservatives-antioxidants (□ = −0.056, p = 0.004), and mixture3 (□ = −0.059, p = 0.043) remained independently associated with reduced Shannon diversity, while NOVA 4 itself was not (□ = −0.006, p = 0.829; Supplementary Figure 4b-c). NOVA 1 (unprocessed foods) showed the expected positive association with diversity (□ = 0.047, p = 0.017; all variance inflation factors < 3.0). This suggests that additive-level resolution may capture additional microbiome variation within UPF consumption that is not reflected by broad NOVA categories.

At the nutrient level, sugar intake from barcoded foods also correlated with both additive variables (e.g., r ≈ 0.5 for TSEGs and pH/AC/L) and reduced gut diversity (r ≈ −0.15; Supplementary Figure 4d), representing a potential confounder. However, in multivariable regression adjusting for sugar intake alongside vegetable and fruit intake, sweeteners-glazing agents remained significantly associated with reduced Shannon diversity (□ = −0.056, p = 0.040), while sugar intake itself was not statistically significant (□ = −0.035, p = 0.088; Supplementary Figure 4e). This suggests that the observed sweetener-glazing agent association is not solely attributable to co-consumed added sugars from processed foods.

Several limitations should be acknowledged. First, additive exposures were estimated using the count of distinct E-codes per barcoded food item, and not additive concentrations. This may lead to over/under-estimation of the influence of the additive, particularly for trace-level additives. Second, barcode scanning accounted for only about 15% of logged meals in Food & You study, with the remainder recorded via photo or manual annotation. Although barcode logging offers high specificity, it may not capture all processed foods consumed, introducing potential selection bias. For instance, alcoholic beverages are exempt from mandatory ingredient listing under EU Regulation 1169/2011, which may result in incomplete additive capture for this food group, though it represented a small share of barcoded food logging in our study. Third, microbiome composition was assessed using a single 16S rRNA sample per participant, which does not capture temporal variability or functional gene content, thus limiting causal relationship investigation. Finally, our cohort consisted of generally healthy, educated Swiss adults, limiting generalizability to populations with different dietary practices or geographies.

Overall, our findings underscore the importance of considering additive exposure in dietary assessments and public health strategies. Food additives are associated with consistent reductions in gut microbiota diversity, even in individuals with otherwise healthful diets. These effects appear to be independent of fast food consumption and are most strongly captured when considered in relation to protective dietary factors. Future dietary guidelines may benefit from incorporating additive reduction alongside promotion of whole, nutrient-dense foods. As UPFs consumption continues to rise globally, recognizing the microbiota as a potential mediator of diet-related health outcomes becomes increasingly critical. Our study provides a framework for integrating additive metrics into microbiota-informed nutrition science and highlights the need for further research on the long-term health implications of additive cocktails in the human diet.

## Methods

### Study design and participants

We analyzed data from the “Food & You” digital nutrition cohort, which enrolled 1,013 healthy adult volunteers in Switzerland between 2019 and March 2023 (Geneva Ethics Commission 2017□02124; SNCTP000002833; NCT03848299). Participants were recruited online and self-screened for eligibility before enrollment. Sociodemographic, behavioral, and anthropometric data were collected via online questionnaires administered during the enrollment phase. Exclusion criteria included current pregnancy, dialysis, use of immunosuppressive medication, antibiotic use in the past 3 months, chronic gastrointestinal disease, active inflammatory or neoplastic disease within the last three years, neuropsychiatric disorders, myocardial infarction or cerebrovascular accident in the past six months, and a prior diagnosis of type 1 or type 2 diabetes mellitus. Of the enrolled participants, 56% were female, 80% were aged 18–49 years, and the majority had normal BMI (67%), with 24% classified as overweight and 7% as obese. Detailed recruitment procedures, study flow and cohort characteristics are reported in Héritier et al. (2023)^18^.

Participants tracked their usual diets for 2–4□weeks under free□living conditions, logging all meals and packaged foods via the AI□assisted MyFoodRepo smartphone app (photo recognition, barcode scan or manual entry). Real-time food logging through photo capture or barcode scanning, combined with automatic image recognition and subsequent validation by expert human annotators, minimized recall bias by eliminating reliance on retrospective reporting, resulting in high adherence rates (>90% over two weeks of tracking) The MyFoodRepo image recognition algorithm has been benchmarked for food classification accuracy^29^, the tool’s dietary assessment performance has been validated against weighted food diaries showing high accuracy for food identification^30^, and cohort-level nutritional patterns showed good agreement with MenuCH, a nationally representative Swiss dietary survey^18^. A single stool sample was collected during the tracking period for 16S rRNA gene sequencing (n□=□992 after rarefaction). Daily energy intakes <□1,000□kcal and participants with ≤5□valid tracking days were excluded, yielding 975 individuals for analyses of diet variability. All participants provided written informed consent.

### Food Additives Data Processing

We obtained additive intake information from users who logged barcoded foods. These barcoded foods were referenced against the OpenFoodFacts database^19^ which provided the E-coded additives (i.e., additives classified under European food safety regulations) present in those foods. In our study additive intake refers to the counts of the additives rather than their amounts since the amount wasn’t available for all foods. Mean daily additive intake per user was calculated by summing the number of unique additives consumed each day and dividing by total tracking days.

Across the cohort, 12,110 unique barcoded food items were logged, of which 4,119 (34%) contained at least one additive. Participants recorded approximately 43,000 barcoded food intake occasions in total, of which 16,300 (38%) included additives.

Barcoded food consumption events containing at least one additive were assigned to food groups and also labeled by day of week based on the recorded consumption timestamp. To examine temporal patterns in consumption of additive-containing food groups, food intake records were aggregated by food group and day of week. For each food group, the total number of intake occasions was computed for each weekday across all participants. For visualization (Figure 1c), weekly profiles were normalized within each food group such that values summed to one across the seven days.

Non□negative matrix factorization of additive mixtures Additive intake patterns were analyzed using non-negative matrix factorization (NMF). This approach has been used to define real-world additive mixtures in NutriNet-Santé and to link two of those mixtures to higher type□2 diabetes risk^15,17^. Additives observed in fewer than 5 participants were excluded, leaving 196 of 257 additives for analysis. NMF models were fitted using scikit-learn^31^ package in Python using coordinate descent and Frobenius norm. The number of components (k) was chosen by maximizing the cophenetic coefficient over k=2 to 10, based on 30 random initializations per k. For each run, participants were assigned to the component with the highest NMF score, a consensus matrix was formed by averaging co-membership matrices across runs, and the cophenetic coefficient was computed via average-linkage hierarchical clustering on 1 minus the consensus matrix. The most stable solution (k=3) was retained (Supplementary Figure 5a).

The final model was fit using NNDSVD initialization (max 5000 iterations, tolerance 1e-4). Component loadings (H) were extracted to quantify each additive’s contribution to each mixture. To define emblematic additives, loadings were row-normalized to represent relative contributions within each mixture, and additives with normalized contribution at least 0.05 in at least one mixture were retained for visualization (Figure 2a). For this heatmap, emblematic additives were hierarchically clustered using average linkage on the raw loading values, while the displayed values corresponded to absolute (raw) loadings. The dominant mixture for each participant was determined by identifying the component with the highest score. Stability of the three-component solution was confirmed through repeated initializations, and co-occurrence of high exposure across mixtures was assessed using conditional probabilities (see Supplementary Methods).

### Microbiome Data Processing

Participants in the “Food & You” study provided a single stool sample during their tracking phase, which were processed following protocols detailed in Singh et al. (2025)^32^. Briefly, the V4 region of the 16S rRNA gene was amplified using 515F and 806R primers and sequenced via two-step Nextera PCR libraries at Microsynth AG (Switzerland). Preprocessing utilized QIIME 2 (version 2024.2)^33^, with demultiplexed single-end reads denoised using Deblur (version 2024.2)^34^ to construct amplicon sequence variants (ASVs) at 150bp. Taxonomic classification was performed against the Greengenes2 database (version 2024.09)^35^. After rarefaction to 15,000 reads per sample, 992 samples were retained for analysis. Alpha diversity metrics included Shannon diversity, Faith’s phylogenetic distance, Pielou’s evenness, and observed features, all computed using q2-diversity (version 2024.2). For detailed methods on microbiome data generation and processing, see Singh et al. (2025)^32^.

### Groupwise association testing

To examine relationships between additives, mixtures and dietary characteristics, we conducted a series of statistical comparisons using non-parametric tests. For categorical variables with multiple groups (e.g., BMI categories, diet quality quartiles), we performed the Kruskal-Wallis (KW) test^36^, while for binary comparisons (e.g., above/below mean threshold, language groups), we used the Mann-Whitney U (MW) test^36^. Statistical significance was set at α=0.05. For multiple group comparisons, we performed post-hoc pairwise tests with appropriate correction. Quartiles were created for continuous variables to facilitate group comparisons. Results were visualized using boxplots with statistical annotation bars indicating significance levels (*p<0.05, **p<0.01, ***p<0.001, ****p<0.0001). For each statistical test, we reported the test statistic (H or U) and corresponding nominal p-value directly on each subplot.

To characterize how additive intake varies across dietary, lifestyle, and demographic characteristics (Figure 1d), we used two complementary approaches depending on predictor type. For continuous predictors, associations with each additive class were quantified using Spearman rank correlations. For categorical predictors, differences in additive exposure were evaluated using two-sided Mann–Whitney U tests, with effect sizes quantified using the rank biserial correlation, where positive values indicate higher exposure in the first listed category. Additive intake variables were log1p transformed prior to analysis. P-values were adjusted for multiple testing using the Benjamini–Hochberg false discovery rate (FDR) procedure, and results are reported as FDR-adjusted q-values (q < 0.05)..

### Beta diversity ordination and environmental fitting

Gut microbiota beta diversity was assessed using unweighted UniFrac distances calculated from rarefied 16S rRNA gene profiles. Principal coordinates analysis (PCoA) was performed on the distance matrix using eigen decomposition, and the first two axes were retained for visualization. Associations between additive intake and beta diversity were tested using PERMANOVA (adonis2, 999 permutations) with additive intake modeled as a continuous variable. Adjusted models included additive intake and the same set of covariates used in the alpha diversity regression analyses. Separate models were fitted for total additive intake, TSEGs, SG, PA, and pH/AC/L. Homogeneity of within-group dispersion was assessed using PERMDISP (betadisper with permutest in R), with participants grouped into quartiles of each additive variable.

Environmental fitting was performed using the envfit function from the vegan package to project dietary and additive variables onto the unconstrained PCoA ordination. Fitted variables included additive classes, additive mixtures, food group intakes (fruits, vegetables, meat, fast food, and alcohol) and HEI. Vectors were visualized as arrows originating from the ordination origin, where arrow direction indicates the gradient of each variable and arrow length reflects the strength of correlation with the ordination axes.

### Regression modeling of microbial diversity

We assessed associations between food additive exposures and gut microbiota diversity, quantified using Shannon diversity, using multivariable regression models specified in statsmodels python package^37^. Covariates were selected a priori based on their potential to confound associations between dietary additive exposure and gut microbiome composition, as they are known to influence both dietary patterns and microbial diversity^32,38,39^. These included demographic characteristics (age, BMI gender), host and behavioral factors (smoking status, alcohol consumption, antibiotic treatment, defecation frequency per day), geographic variables (language region and urbanity), dietary factors (HEI and total energy intake), and sequencing batch. For mixture analyses, we fit an ordinary least squares (OLS) regression model including the three mixture variables (mixture1, mixture2, mixture3) as primary predictors and adjusting for aforementioned covariates (n = 940 after excluding observations with missing data for language region and urbanity).

For additive class analyses, namely SG and PA, we fit analogous covariate adjusted OLS models in which the mixture terms were replaced by the additive class. Inference for these OLS models used heteroskedasticity consistent (HC3) standard errors.

The regression model can be represented by the following equation:

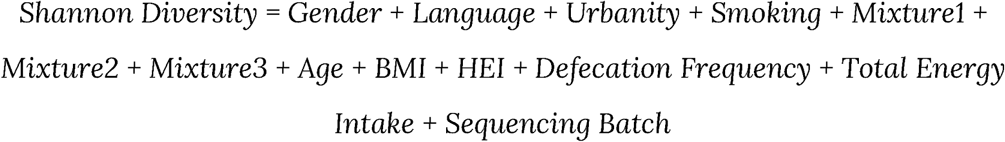

To validate model assumptions and ensure the reliability of the regression estimates, we performed several diagnostic tests. The Breusch–Pagan test applied to residuals from the standard OLS model indicated no evidence of heteroskedasticity (Lagrange multiplier p = 0.94; F test p = 0.94). Multicollinearity was assessed using variance inflation factors (VIF), with all predictors showing low collinearity (all VIF < 3). To assess robustness of mixture associations to influential observations and deviations from OLS assumptions, we additionally fit robust linear regression models using Huber loss (RLM with HuberT norm). Robust model estimates were further evaluated using nonparametric bootstrap resampling (1000 bootstrap samples, sampling participants with replacement), providing an additional sensitivity analysis for mixture effect estimates.

To examine the relationships between dietary factors and gut microbiome diversity, we conducted multivariable linear regression analyses using OLS with Shannon entropy as the dependent variable (Figure 3). All continuous dietary variables (vegetable intake, fruit intake, fast food consumption, and total additive intake) were standardized (z-scored) by subtracting the mean and dividing by the standard deviation to enable direct comparison of effect sizes across different measurement scales.

We fitted three separate models to test specific dietary interactions: (1) vegetable intake × total additive intake, (2) fruit intake × total additive intake, and (3) fast food consumption × total additive intake, with the latter including standardized vegetable intake as a covariate. Each model included the same set of covariates: age, BMI, gender, urbanity, Swiss linguistic region, smoking status, alcohol consumption, antibiotic treatment history, defecation frequency, total energy intake, and sequencing batch. HEI was excluded from these models to avoid collinearity, as vegetable and fruit intakes are direct components of the HEI score. Interaction terms were constructed by multiplying the standardized main effect variables. All models used heteroscedasticity-robust standard errors (HC3) to account for potential non-constant variance.

In addition to total additive intake, separate models were estimated for each additive class (e.g., colorants, sweeteners, preservatives, emulsifiers, mixtures). Interaction coefficients and confidence intervals from these models were extracted to compare the extent to which different additives attenuated the vegetable–diversity association.

Regression modeling outputs, including coefficient estimates, confidence intervals, and model diagnostics for all regression analyses, are provided in Supplementary File 1. Nominal p-values are reported for all regression models, as each model tests a limited number of pre-specified primary predictors.

### Dietary contrast metrics

To examine the balance between health-associated dietary components and additive or fast-food intake, we derived composite ‘minus’ metrics by subtracting the z-score of each additive class or fast-food variable from the z-score of a reference dietary variable (HEI, fruits, or vegetables). For example, HEI-minus-PA represents the standardized difference between HEI diet quality and preservative/antioxidant exposure. Positive values indicate relative predominance of the health-associated component; negative values indicate relative predominance of the additive or fast-food component.

### Differential Abundance Analysis using BIRDMAn

To identify taxa associated with dietary patterns, we performed differential abundance analysis using BIRDMAn (Bayesian Inferential Regression for Differential Microbiome Analysis)^20^. Raw feature count data were filtered to retain features present in at least 25% of samples. For each exposure, we included the exposure of interest and sequencing batch as a fixed effect covariate. Models were fit using variational inference with 500 posterior draws. Taxa were considered credibly associated with the exposure if the highest density interval did not include zero, and effect sizes were summarized as the posterior mean.

For each additive category and food group, we first selected up to the top 100 ASVs ranked by absolute posterior mean effect size whose 95% highest density interval for the effect excluded zero. Among these credibly associated ASVs, we defined the numerator as the set with positive posterior mean effect size and the denominator as the set with negative posterior mean effect size. For each sample, we summed raw counts across numerator ASVs and across denominator ASVs, excluded samples with zero total counts in either sum, and computed the log10 ratio of the numerator sum to the denominator sum. All log-ratio metrics were correlated with nutritional variables using Pearson’s r.

To identify microbial taxa consistently associated with additive exposures, we extracted mean posterior effect sizes from BIRDMAn differential abundance models. We first defined a union set of ASVs appearing in the numerator or denominator of log-ratio balances for TSEGs, SG, and pH/AC/L, capturing taxa commonly modulated across these additive categories. For each dietary variable (additive-only and vegetables-minus contrasts), the top 50 ASVs by absolute effect size were retained and filtered to include only those present in this union set. To focus on taxa with robust additive associations, we retained ASVs whose summed absolute effect sizes across the four additive-only variables exceeded 0.25. The complete set of differentially abundant taxa with sequences for each dietary variable is provided in Supplementary Table 4.

To visualize differential responses across taxa and exposures, a scatter-style heatmap was constructed. Features were hierarchically clustered (Ward’s method, Euclidean distance) based on their effect size profiles, and sorted accordingly. Each dot represents a microbial feature’s mean BIRDMAn effect size under a specific exposure, colored along a divergent red–white–blue scale. This enabled comparison of taxa directionality and magnitude across additive and HEI-minus variables, revealing distinct response modules.

### PICRUSt predicted functional pathway analysis

Predicted microbial functional profiles were generated using PICRUSt2^23^, and unstratified pathway abundance tables were used for downstream analyses. Pathways were filtered to retain features present in at least 5% of samples and with non zero variance. Associations between predicted pathway abundances and dietary additive exposures were assessed using multivariable linear models implemented in MaAsLin2 (v1.14.1)^40^. Models were fitted separately for each additive exposure and dietary mixture, with pathway abundance as the outcome and the additive exposure of interest included as a fixed effect. Each model adjusted for age, BMI, gender, language region, urbanity, antibiotic treatment status, smoking status, defecation frequency per day, and sequencing batch. Pathway abundances were normalized using total sum scaling and log transformed within MaAsLin2.

Multiple testing correction was performed using the Benjamini–Hochberg false discovery rate (FDR) procedure across all pathway–exposure associations, and results are reported as FDR-adjusted q-values (q < 0.05). For Figures 5c-d, complementary OLS models with heteroscedasticity robust standard errors were additionally fitted using log transformed pathway abundances, and FDR corrected results were used for labeling and cross validation of pathway level associations (Supplementary Table 3). Associations between pathway abundances significantly associated with additive features (SG and mixture3) and Shannon diversity were evaluated using Pearson correlation.

### Machine□learning prediction of dietary exposures

To assess whether gut microbial features discriminate between low and high intake of diet components, we trained an XGBoost^41^ classifier on species-level count data. Metadata values for each dietary variable (e.g. HEI, fruits, vegetables) were first dichotomized into low (≤□25th percentile) and high (≥□75th percentile) groups. For each variable, we performed 50 iterations of random stratified splits (80% train, 20% test; random_state□=□0-49). Classifier hyperparameters were fixed at 1,000 trees, max_depth□=□6, η□=□0.005, subsample□=□0.4, colsample_bytree□=□0.8. We recorded area under the receiver-operating-characteristic curve (AUROC) and area under the precision–recall curve (AUPRC) for each iteration, and averaged feature-importance scores across runs to identify the top□40 discriminative taxa. Results were shown in Supplementary File 2 for AUROC/AUPRC and top-ranked features.

To assess how well gut microbial taxa predict continuous measures of diet, we also trained XGBoost regressors on species-level abundance data for each outcome: HEI, fruits intake, vegetables intake, fast-food intake, individual additive classes, and additive/mixture composites. Target variables with very low mean values (<0.001) were log-transformed (log10(x + 1e-6)) to improve model performance. For each target, we performed 50 iterations of random splits without stratification (80% training, 20% testing; random_state = 0–49), using XGBoost with 1,000 boosting rounds and a learning rate of 0.05 to minimize squared-error loss (reg objective). In each iteration, Spearman’s ρ was calculated between observed and predicted values on the held-out test set. The Spearman correlations across iterations are reported in Supplementary File 2.

To explore synergistic effects of additive classes and mixture supplements, we generated all combinations of five selected additives and three mixtures. For each combination, we summed the relevant metadata columns to create a composite exposure variable, then split into low/high as above. Using the same XGBoost-classification pipeline (50 iterations, identical hyperparameters), we computed the mean AUROC for each combination. Top-performing additive and mixture combinations were identified by ranking mean AUROC across all tested sets.

## Data Availability

The sequence data from this study has been deposited in the European Nucleotide Archive (ENA) under the project accession PRJEB85942 and in Qiita under study ID 15880. Metadata containing clinical, demographic, and nutritional variables cannot be deposited publicly due to participant privacy and ethical restrictions. Access to this metadata can be requested by contacting the corresponding author, subject to institutional ethical compliance.

## Code Availability

The analysis code used to generate the results is publicly available on GitHub (https://github.com/digitalepidemiologylab/food-additives-gut-microbiota-public).

## Contributions

R.S.: Conceptualization, data analysis, methodology, visualization, software, investigation, writing - original draft. M.S. and D.M.: Study design, supervision, writing - review and editing. R.K.: Writing - review and editing

## Supplementary Section

### Food group classification

Food items recorded via the MyFoodRepo app were classified into 15 predefined food groups based on their item descriptions: vegetables (including legumes, potatoes, salads, sauces, spices), fruits (including fruit juices), vegan products, meat (including fish, poultry, eggs, processed meat), bread, grains and cereals, dairy, sugary foods (including sweets, soft drinks, chocolate, desserts, pastries), tea, coffee, water, alcohol, oils and nuts, fast food, and others. The fast food category comprised items such as pizza, hamburgers, salty snacks, and other ready-to-eat convenience foods typically associated with quick-service preparation. Food group assignment was performed by matching each food item description to the most appropriate group from the predefined list, with items assigned to ‘others’ only when no suitable group could be determined.

Dietary regularity for food groups, i.e., sugary foods, were quantified using the coefficient of variation (CV), calculated as the standard deviation of daily sugary food intake divided by the mean daily intake across each participant’s tracking period. Higher CV values indicate more irregular day-to-day consumption patterns.

### Mixtures Co-occurrence Analysis

To understand the relationship between high exposure to different additive mixtures, we created a co-occurrence matrix based on binary indicators of high mixture exposure. For each of the three NMF mixture components, participants were classified as having “high” exposure if their score for that mixture was in the top quartile of the distribution. Using these binary indicators, we constructed a co-occurrence matrix by computing the matrix product of the transposed binary matrix with itself. To obtain conditional probabilities, which represent the probability of high exposure to one mixture given high exposure to another, we normalized the co-occurrence matrix by its diagonal elements. This normalization allowed us to assess whether high exposure to certain mixtures tended to occur together in the same individuals.

Co-occurrence analysis of the mixtures revealed a moderate degree of overlap between high additive-intake patterns (Supplementary Figure 5b). Participants with high mixture 1 exposure had a 48 % probability of also scoring high on mixture 3 and 32 % on mixture 2, while those high in mixture 3 showed a 47 % chance of high mixture 1 and 37 % chance of high mixture 2. In contrast, individuals high in mixture 2 had a 32 % probability of high mixture 1 and 38 % probability of high mixture 3. The strongest overlap was observed between mixtures 1 and 3, whereas the mixture 1–2 pairing exhibited the weakest co-occurrence.

### Mixtures Stability Analysis

To evaluate robustness of the selected NMF solution (k=3), we performed a stability analysis based on 50 independent NMF runs with different random initializations. Component loadings were compared across runs using a custom intra-mixture consistency metric. For each mixture, we calculated the average normalized distance between loadings from individual runs and the centroid across all runs, and converted this distance into a stability score using 1/(1+normalized_distance), with values closer to 1 indicating higher stability. The overall stability score of the three-component solution was 1.00, and all individual mixtures showed similarly high stability (mixture1 0.99, mixture2 0.99, mixture3 0.99), indicating highly reproducible additive patterns across random initializations.

---

**Supplementary Figure 1.**
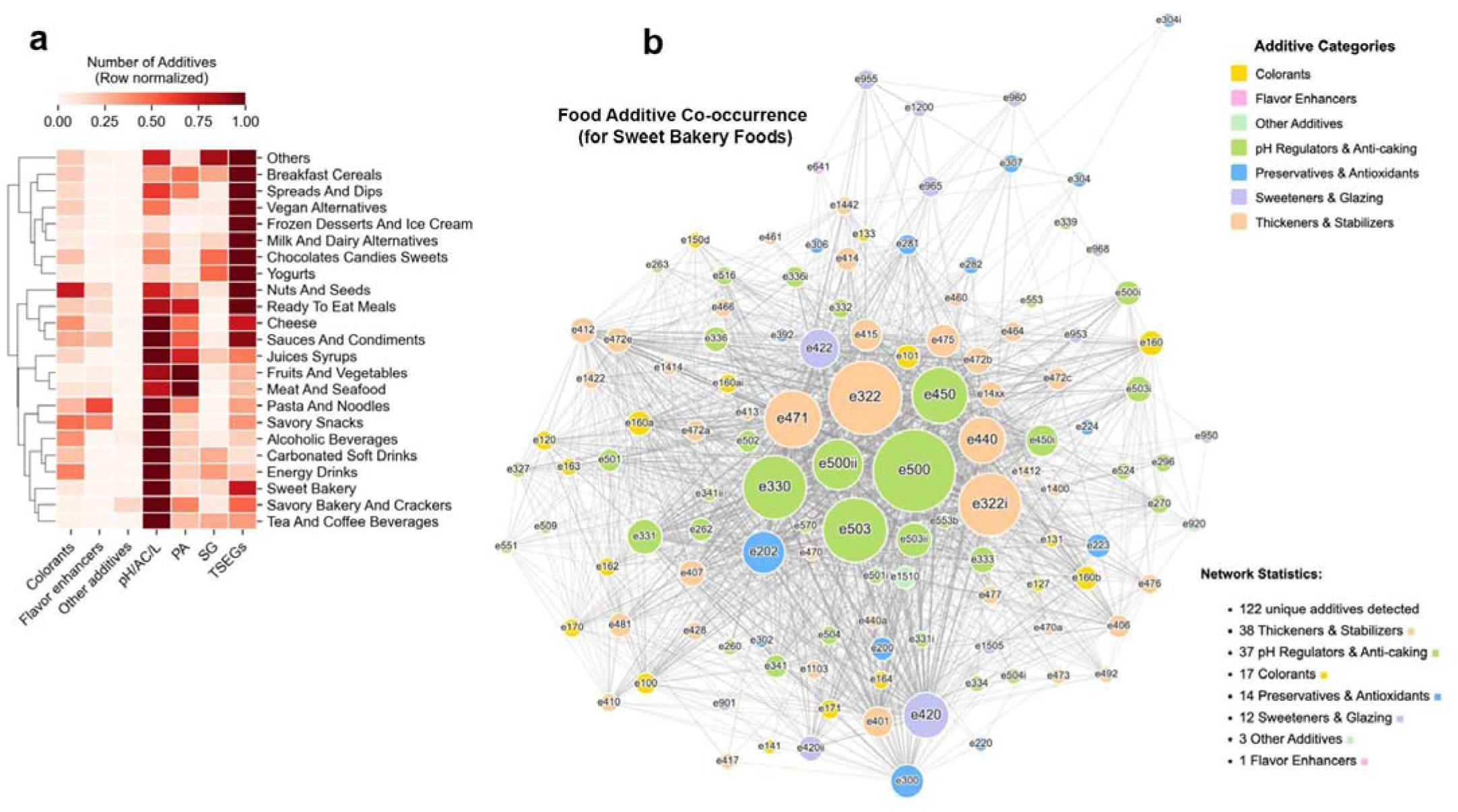
Additive category distribution across food groups and within-product co-occurrence network. **(a)** Distribution of additive categories across food groups. Heatmap showing the relative abundance of each additive category within food groups. Values represent the total count of additives per category aggregated across all unique products within each food group, row-normalized to highlight the proportional composition of additive types within each food group. Food groups (rows) are hierarchically clustered by similarity in additive category profiles; additive categories (columns) are ordered without clustering. Color intensity indicates relative abundance within each row, with darker shades representing higher proportions. **(b)** Co-occurrence network of food additives in sweet bakery products. Each node represents a unique food additive, with node size proportional to the frequency of occurrence within the sweet bakery food group and node color indicating the additive category. Edges connect additives that appear together within the same product, with edge thickness reflecting the frequency of co-occurrence. Nodes are positioned using a force-directed layout algorithm, where additives that frequently co-occur are drawn closer together. Node labels display E-codes for identification.

**Supplementary Figure 2.**
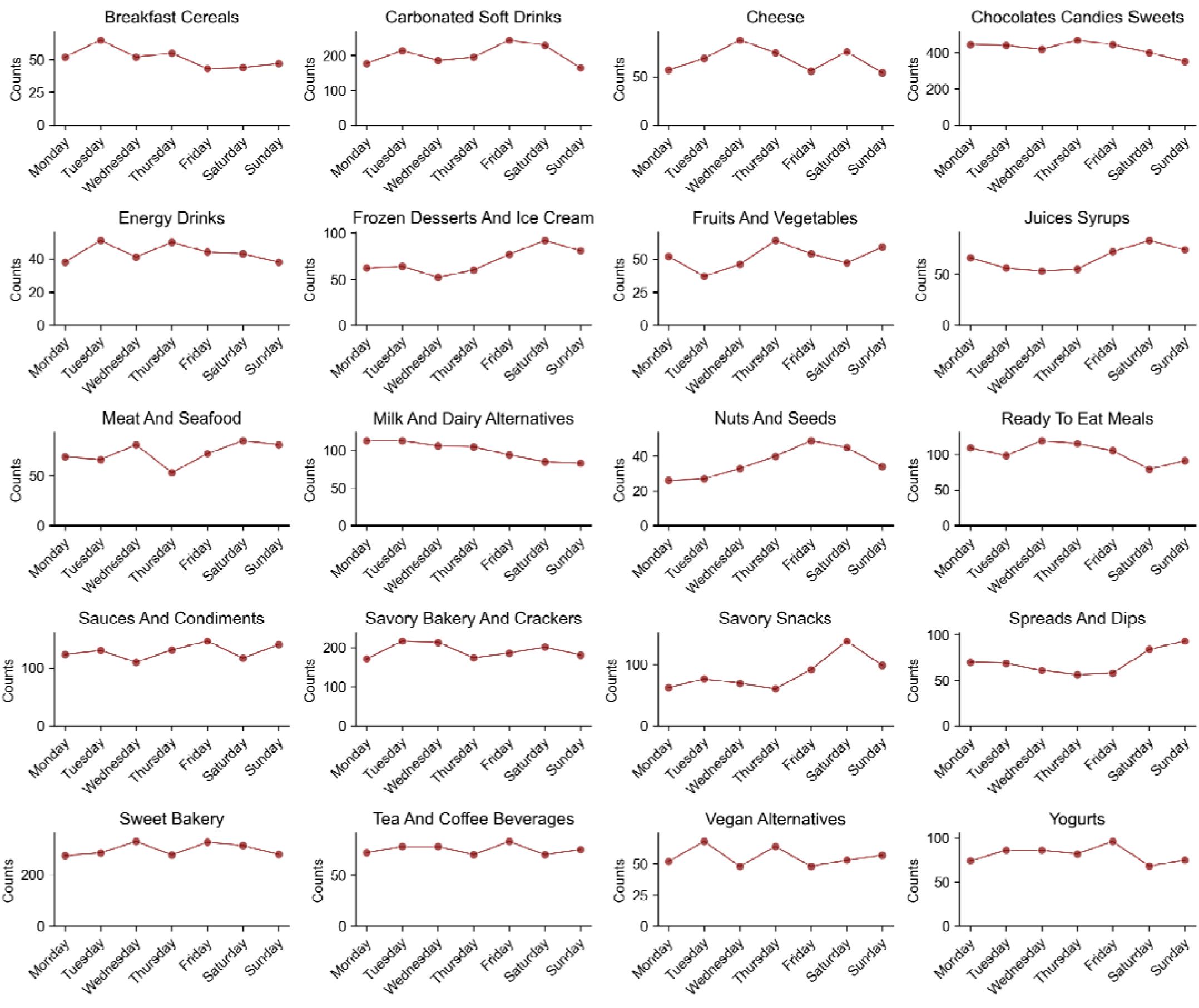
Weekly consumption patterns of additive-containing food groups. Line plots show the distribution of daily consumption counts across days of the week for each food group containing at least one additive.

**Supplementary Figure 3:**
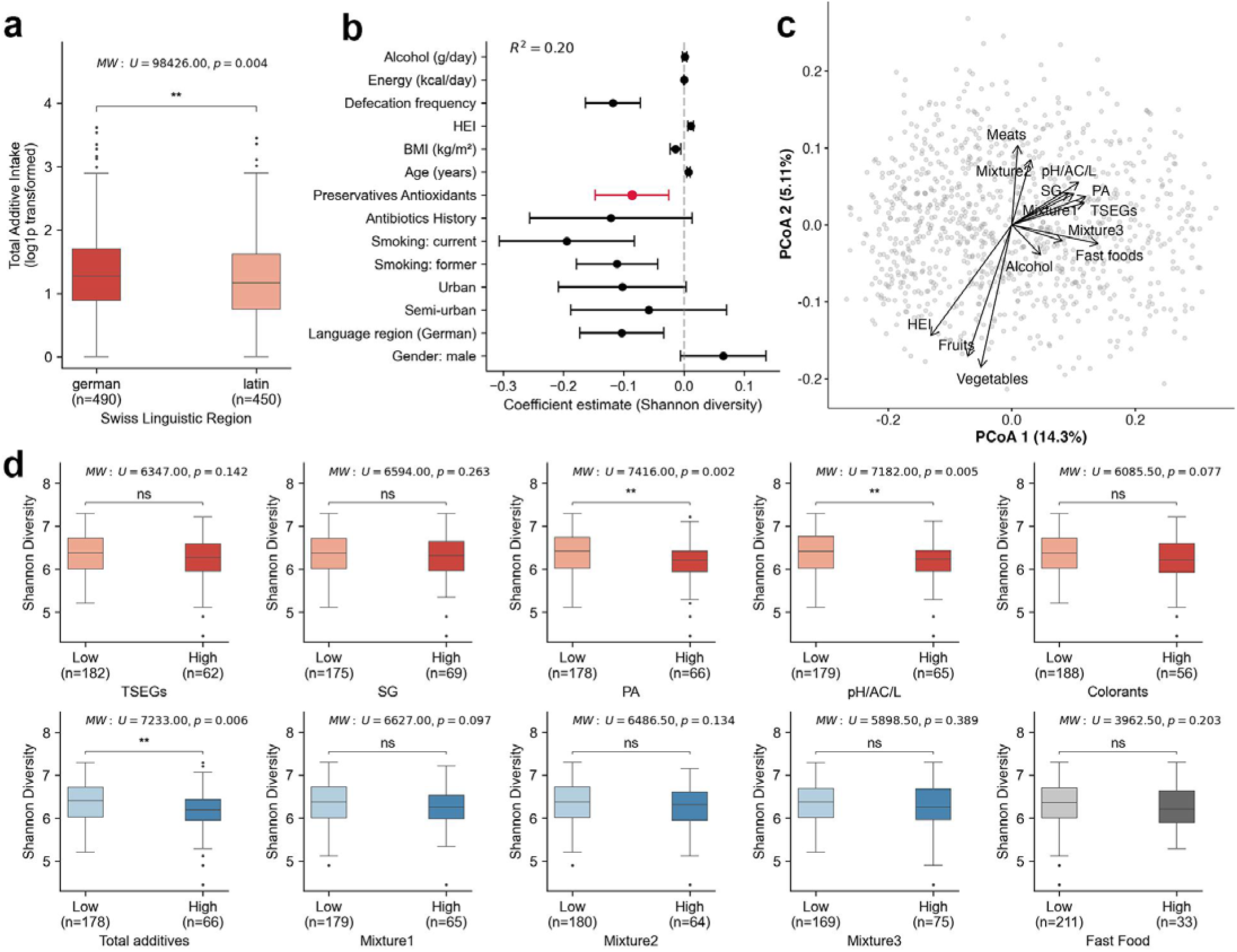
Associations between regional, lifestyle, and dietary factors with additive intake and gut microbial diversity. **(a)** Boxplots comparing total additive intake (log1p transformed) across German-speaking and French/Italian-speaking (Latin) regions of Switzerland. **(b)** Multivariable regression coefficients for the association between preservatives-antioxidants additive intake and gut Shannon diversity (n = 940). Points represent coefficient estimates and horizontal bars indicate 95% confidence intervals. The additive term is highlighted in red. **(c)** Environmental fitting of additive classes, additive mixtures, and dietary variables onto the same PCoA ordination. Arrows originate at the ordination origin and indicate the direction of association of each variable with microbiota composition. **(d)** Impact of concurrent high additive intake on gut diversity among high fruit consumers (>75^th^ percentile). Boxplots compare Shannon diversity between high fruit consumers with low versus high intake of various additives and mixtures. Top row (red) shows individual additive categories. Bottom row shows colorants, total additive intake, additive mixtures and fast food intake. Group differences were assessed using Mann-Whitney U test (panels a and d) and Kruskal-Wallis test with Dunn’s post-hoc comparisons (panel b). Test statistics and p-values are displayed above each plot, with significance indicated (*p <; 0.05, **p <; 0.01, ***p <; 0.001, ****p <; 0.0001, ns = not significant). Sample sizes for each group are shown in parentheses.

**Supplementary Figure 4.**
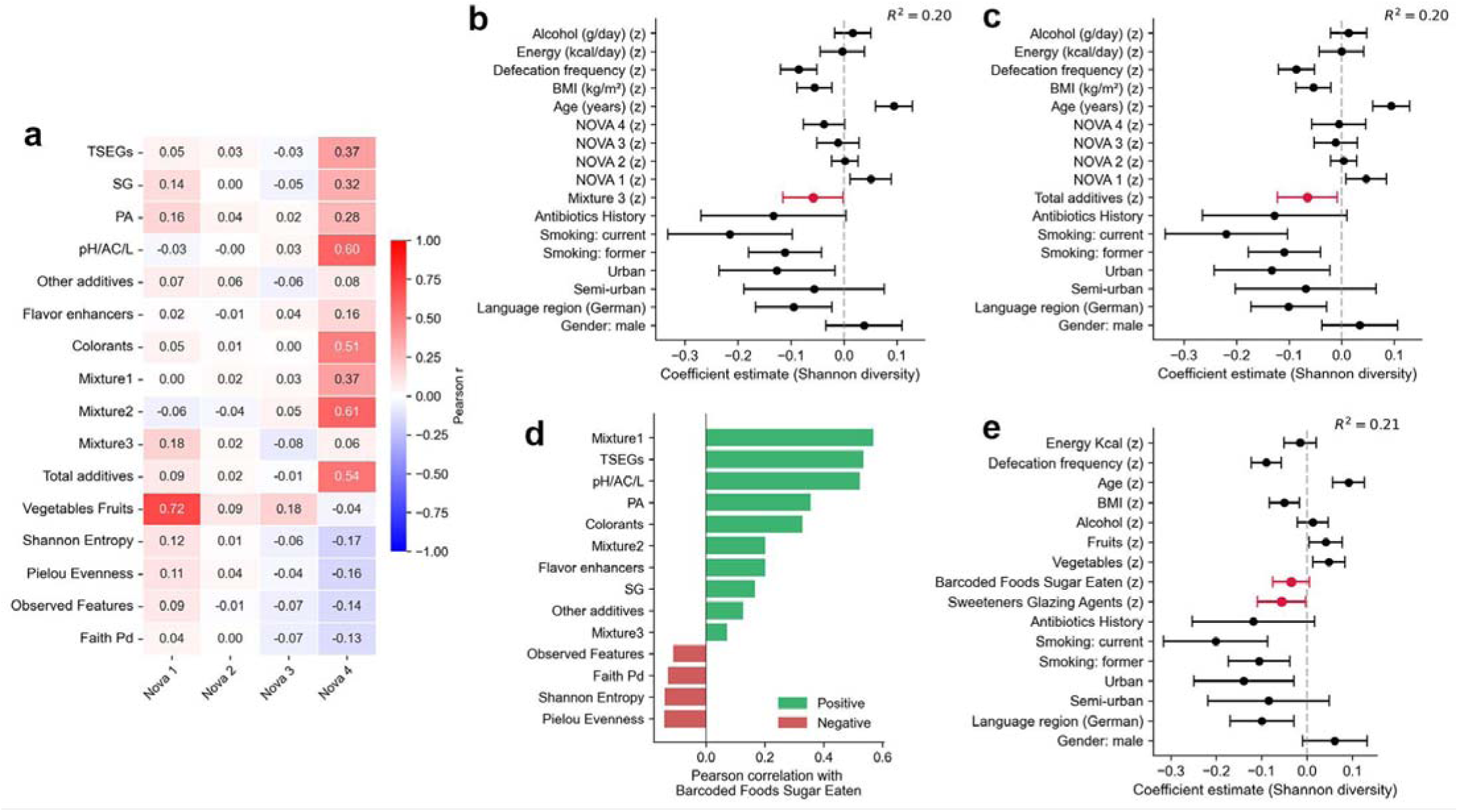
Sensitivity analyses for NOVA food processing categories, sugar intake from barcoded foods, and gut microbiota diversity. **(a)** Pearson correlation heatmap between NOVA food processing categories (NOVA 1–4) and food additive classes, additive mixtures, vegetable and fruit intake, and gut alpha diversity metrics. **(b–c)** Forest plots of multivariable linear regression coefficients predicting gut microbiota Shannon diversity, adjusted for all four NOVA processing categories alongwith demographic, lifestyle, and technical covariates. Panel (b) includes mixture3 and panel (c) includes total additive intake as the primary additive exposure variable (highlighted in red). **(d)** Pearson correlations between sugar intake from barcoded foods and additive classes, additive mixtures, and gut alpha diversity metrics. Green bars indicate positive correlations and red bars indicate negative correlations. **(e)** Forest plot of multivariable linear regression coefficients predicting Shannon diversity, adjusted for sugar intake from barcoded foods, vegetable and fruit intake, and demographic, lifestyle, and technical covariates, with sweeteners-glazing agents as the primary exposure variable (highlighted in red). For all forest plots, points represent coefficient estimates and horizontal bars indicate 95% confidence intervals. Continuous variables are z-standardized; coefficients represent the change in Shannon diversity per one standard deviation increase.

**Supplementary Figure 5:**
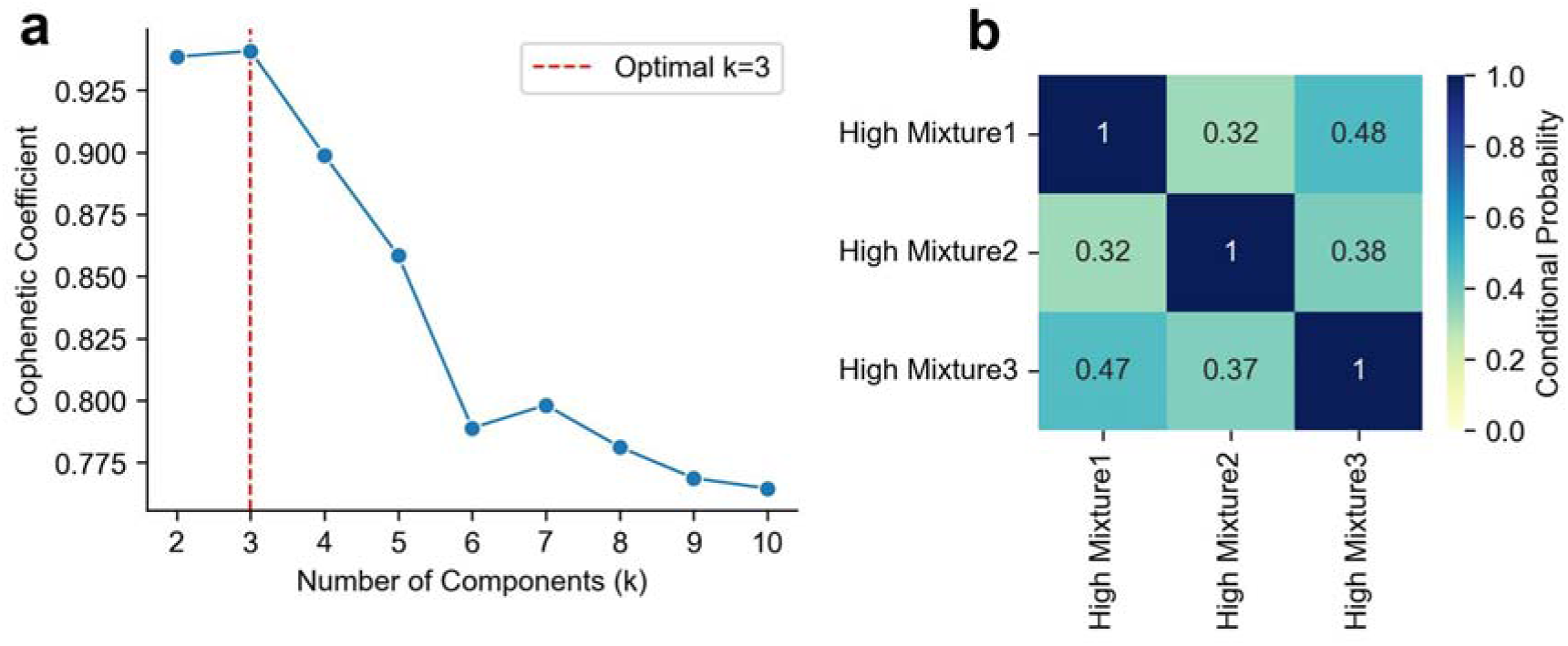
Evaluation of NMF components and co-occurrence patterns. **(a)** Cophenetic coefficient as a function of the number of components (k) for the NMF solution. The line plot shows cophenetic coefficients from k = 2 to 10, with the red dashed vertical line marking the optimal k = 3, highlighting the peak stability at the three-component solution. **(b)** Heatmap of conditional probabilities of high mixture scores. Each cell shows P(high Mixture j | high Mixture i), calculated by normalizing co-occurrence counts by the total number scoring high on the row pattern. Moderate overlap is evident, especially between mixtures 1 and 3.

## Supplementary Tables

**Supplementary Table 1. Prevalence of food additives across study participants.**

**Supplementary Table 2. Non-negative matrix factorization (NMF) loadings for dietary additive mixtures. Each row represents a food additive (identified by E-code and name), and each column represents an identified additive mixture pattern. Values indicate the loading coefficients (contribution weights) of each additive to each mixture, with higher values indicating stronger association with that mixture pattern. Only additives with loadings ≥0.05 in at least one mixture are included.**

**Supplementary Table 3. Food additive and mixture associations with microbial predicted pathways from PICRUSt2.**

**Supplementary Table 4. BIRDMAn effect size estimates and ASV sequences for taxa associated with dietary and additive variables.**

**Supplementary File 1. Regression modeling outputs for gut microbiota Shannon diversity analyses.**

**Supplementary File 2. XGBoost modeling results for classification and regression tasks.**

