## Supplementary for "Food additive exposure associated with reduction in gut microbiota diversity"

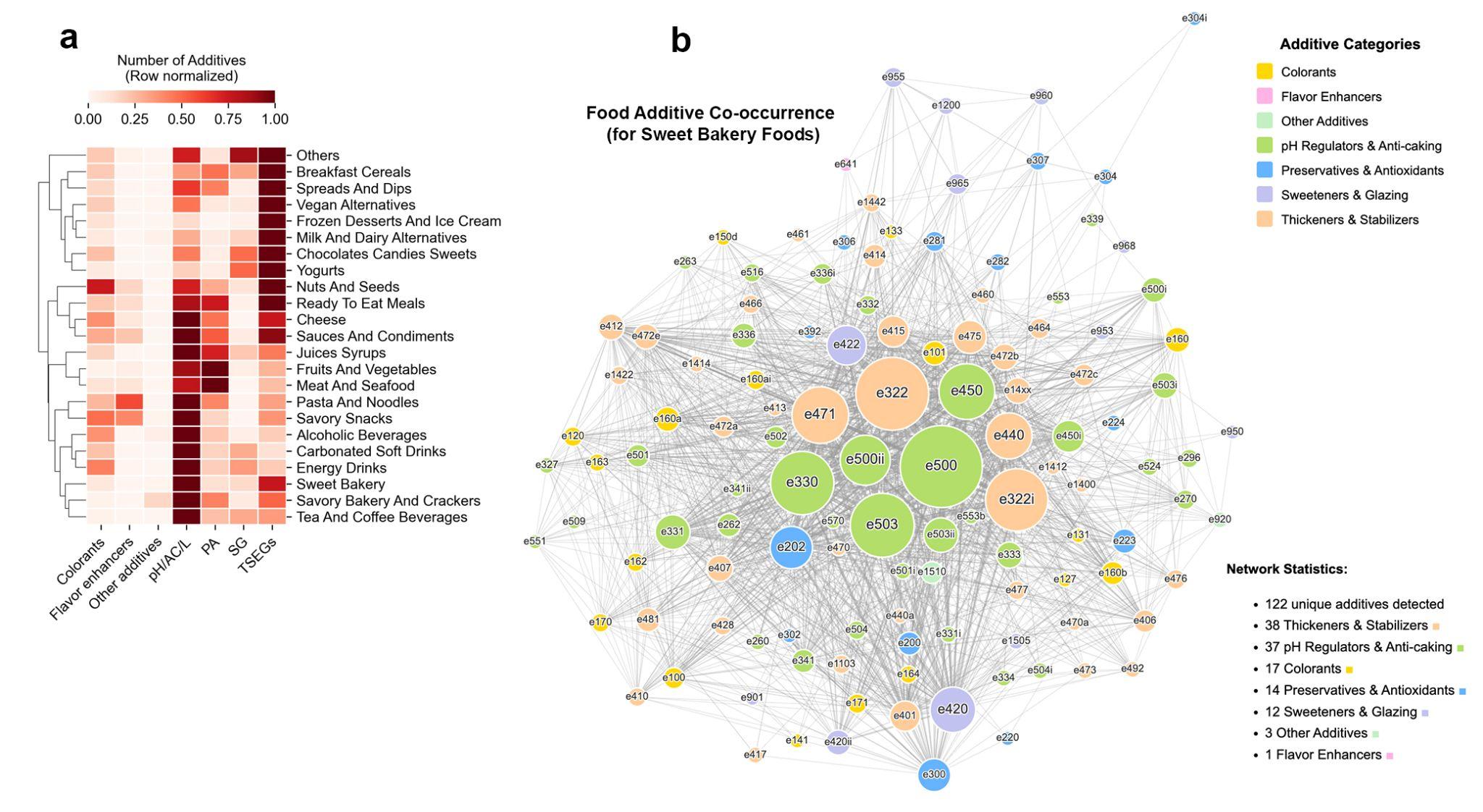

**Supplementary Figure 1. Additive category distribution across food groups and within-product co-occurrence network.** **(a)** Distribution of additive categories across food groups. Heatmap showing the relative abundance of each additive category within food groups. Values represent the total count of additives per category aggregated across all unique products within each food group, row-normalized to highlight the proportional composition of additive types within each food group. Food groups (rows) are hierarchically clustered by similarity in additive category profiles; additive categories (columns) are ordered without clustering. Color intensity indicates relative abundance within each row, with darker shades representing higher proportions. **(b)** Co-occurrence network of food additives in sweet bakery products. Each node represents a unique food additive, with node size proportional to the frequency of occurrence within the sweet bakery food group and node color indicating the additive category. Edges connect additives that appear together within the same product, with edge thickness reflecting the frequency of co-occurrence. Nodes are positioned using a force-directed layout algorithm, where additives that frequently co-occur are drawn closer together. Node labels display E-codes for identification.

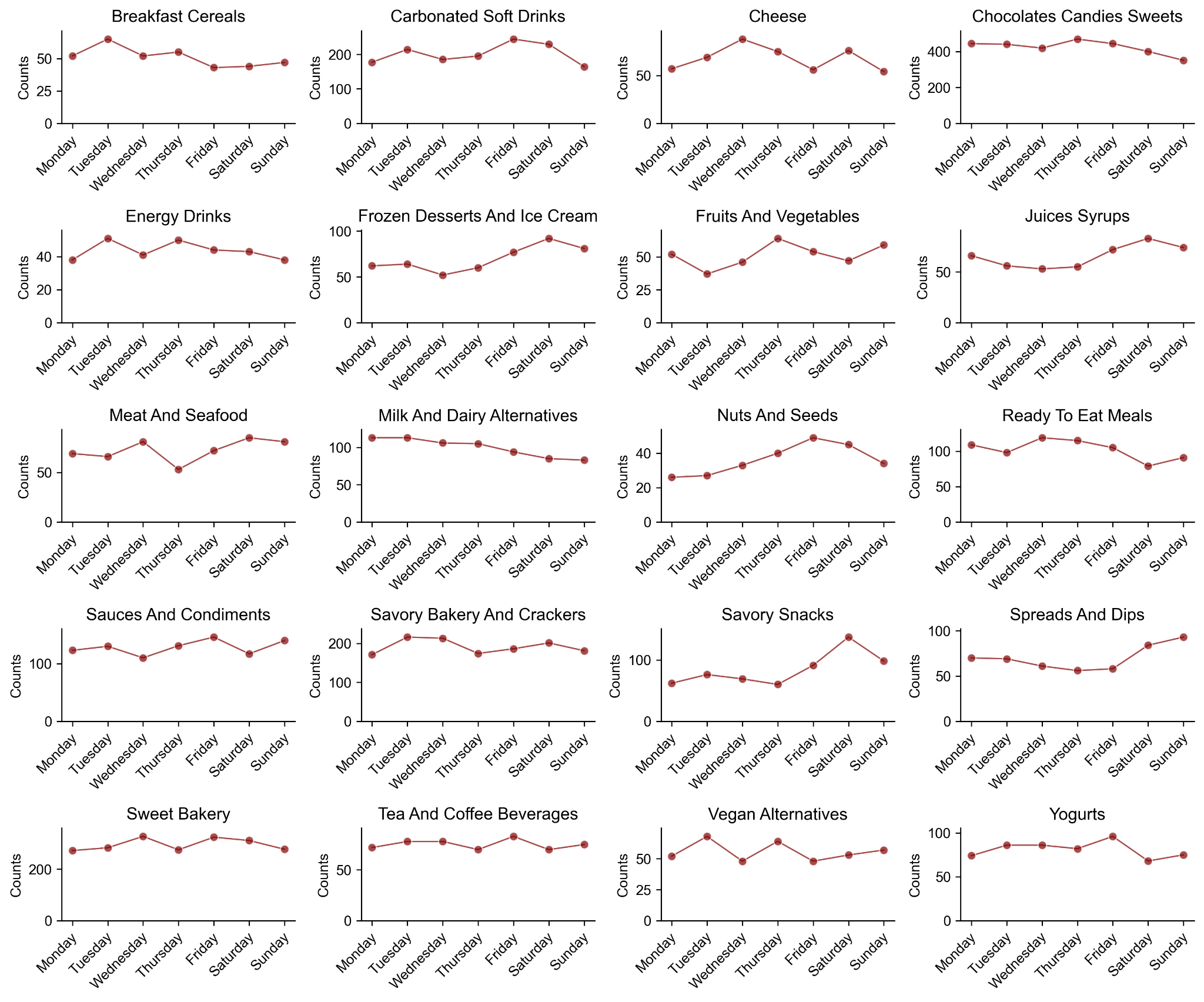

**Supplementary Figure 2. Weekly consumption patterns of additive-containing food groups.** Line plots show the distribution of daily consumption counts across days of the week for each food group containing at least one additive.

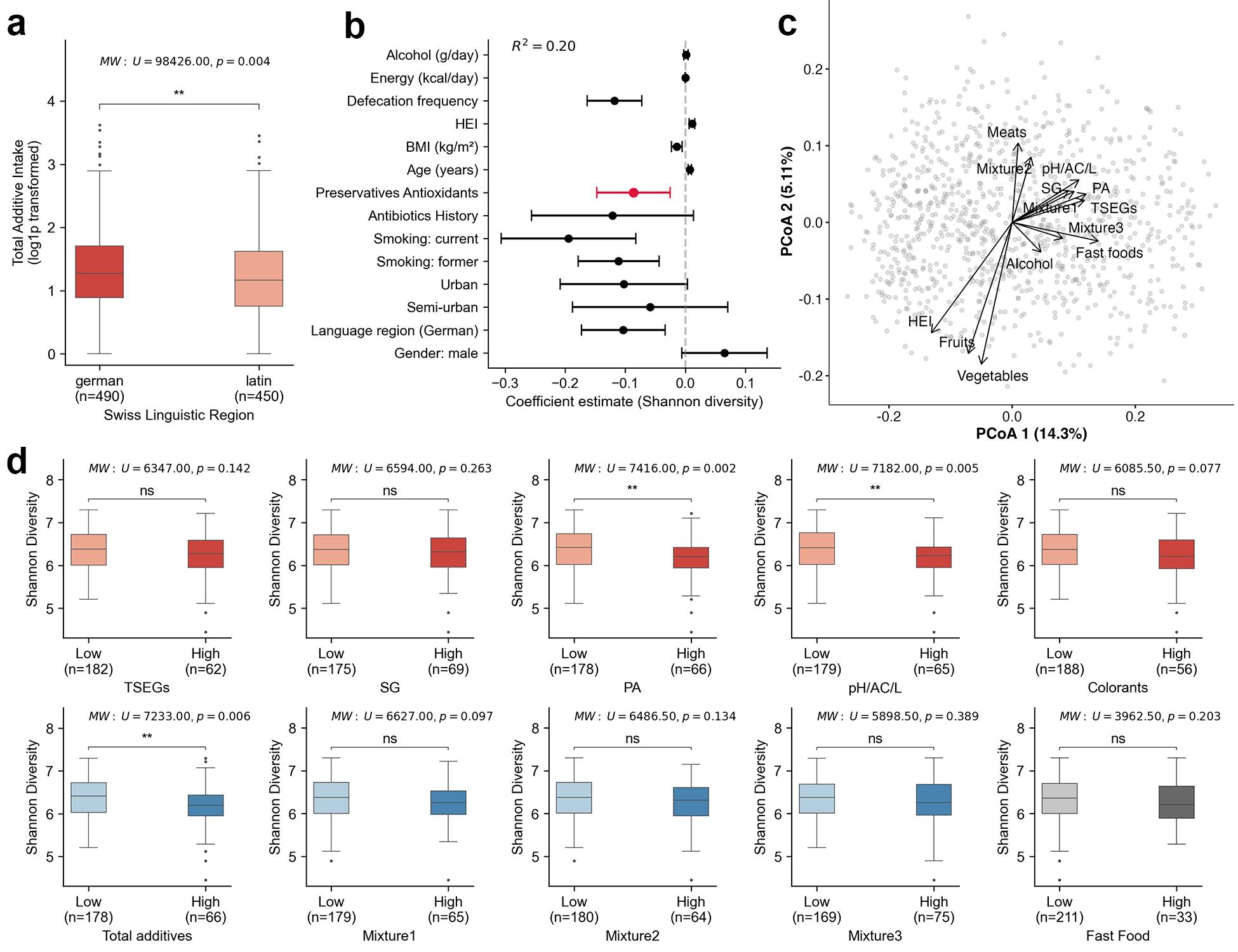

**Supplementary Figure 3: Associations between regional, lifestyle, and dietary factors with additive intake and gut microbial diversity. (a)** Boxplots comparing total additive intake (log1p transformed) across German-speaking and French/Italian-speaking (Latin) regions of Switzerland. **(b)** Multivariable regression coefficients for the association between preservatives-antioxidants additive intake and gut Shannon diversity (*n* = 940). Points represent coefficient estimates and horizontal bars indicate 95% confidence intervals. The additive term is highlighted in red. **(c)** Environmental fitting of additive classes, additive mixtures, and dietary variables onto the same PCoA ordination. Arrows originate at the ordination origin and indicate the direction of association of each variable with microbiota composition. **(d)** Impact of concurrent high additive intake on gut diversity among high fruit consumers (>75^th^ percentile). Boxplots compare Shannon diversity between high fruit consumers with low versus high intake of various additives and mixtures. Top row (red) shows individual additive categories. Bottom row shows colorants, total additive intake, additive mixtures and fast food intake. Group differences were assessed using Mann-Whitney U test (panels a and d) and Kruskal-Wallis test with Dunn's post-hoc comparisons (panel b). Test statistics and p-values are displayed above each plot, with significance indicated (*p < 0.05, **p < 0.01, ***p < 0.001, ****p < 0.0001, ns = not significant). Sample sizes for each group are shown in parentheses.

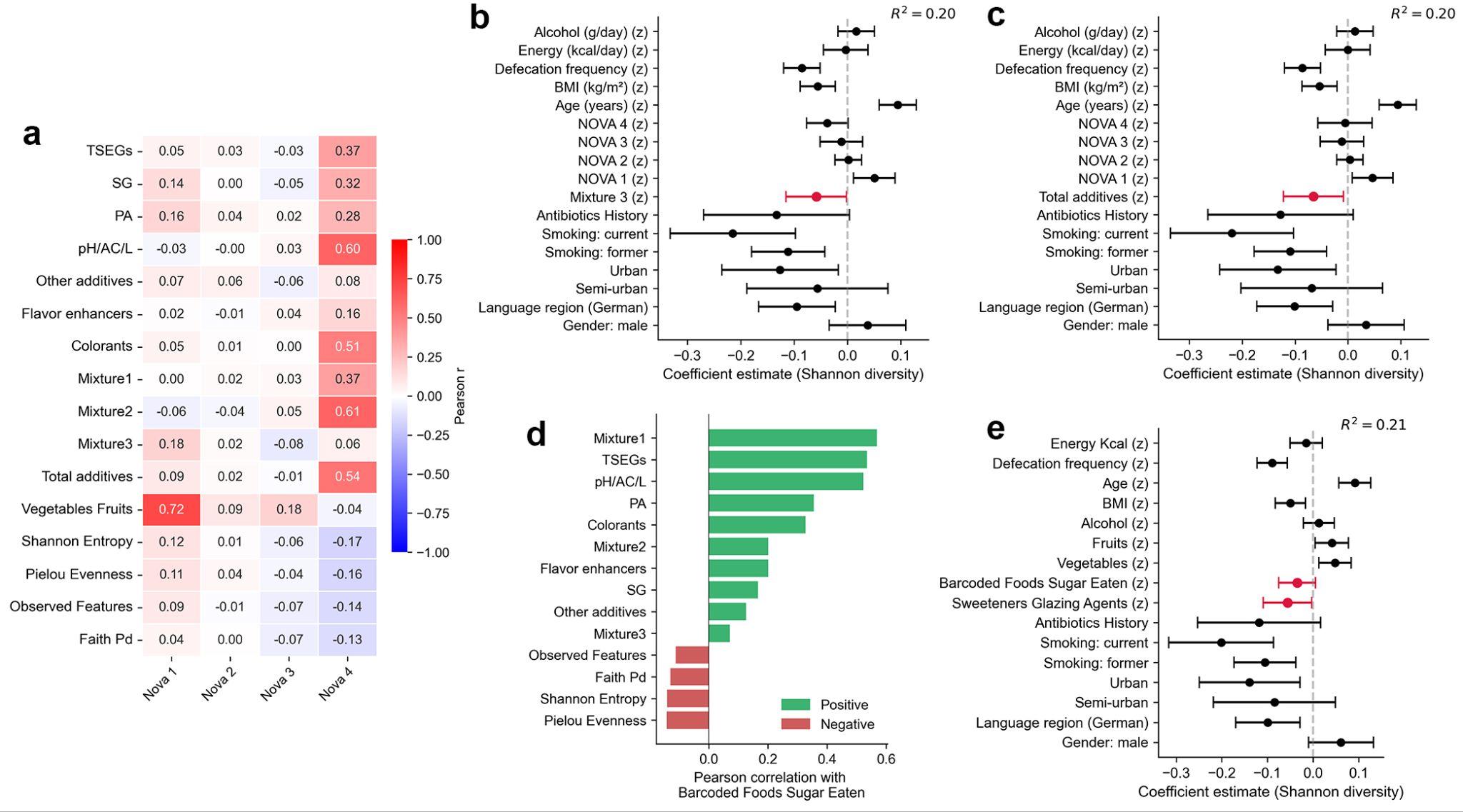

**Supplementary Figure 4. Sensitivity analyses for NOVA food processing categories, sugar intake from barcoded foods, and gut microbiota diversity. (a)** Pearson correlation heatmap between NOVA food processing categories (NOVA 1–4) and food additive classes, additive mixtures, vegetable and fruit intake, and gut alpha diversity metrics. **(b–c)** Forest plots of multivariable linear regression coefficients predicting gut microbiota Shannon diversity, adjusted for all four NOVA processing categories alongwith demographic, lifestyle, and technical covariates. Panel (b) includes mixture3 and panel (c) includes total additive intake as the primary additive exposure variable (highlighted in red). **(d)** Pearson correlations between sugar intake from barcoded foods and additive classes, additive mixtures, and gut alpha diversity metrics. Green bars indicate positive correlations and red bars indicate negative correlations. **(e)** Forest plot of multivariable linear regression coefficients predicting Shannon diversity, adjusted for sugar intake from barcoded foods, vegetable and fruit intake, and demographic, lifestyle, and technical covariates, with sweeteners-glazing agents as the primary exposure variable (highlighted in red). For all forest plots, points represent coefficient estimates and horizontal bars indicate 95% confidence intervals. Continuous variables are z-standardized; coefficients represent the change in Shannon diversity per one standard deviation increase.

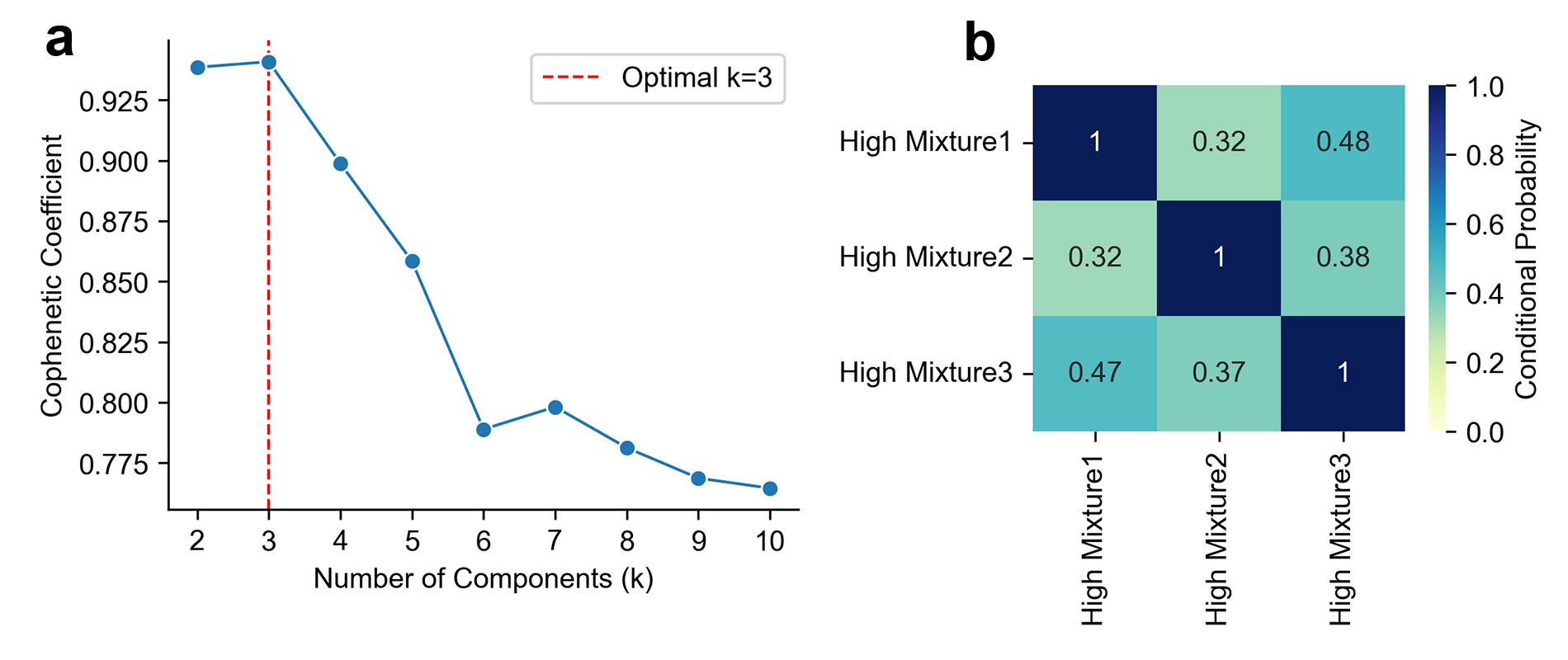

**Supplementary Figure 5: Evaluation of NMF components and co-occurrence patterns.** **(a)** Cophenetic coefficient as a function of the number of components (k) for the NMF solution. The line plot shows cophenetic coefficients from k = 2 to 10, with the red dashed vertical line marking the optimal k = 3, highlighting the peak stability at the three-component solution. **(b)** Heatmap of conditional probabilities of high mixture scores. Each cell shows P(high Mixture j | high Mixture i), calculated by normalizing co-occurrence counts by the total number scoring high on the row pattern. Moderate overlap is evident, especially between mixtures 1 and 3.

### Supplementary Tables

**Supplementary Table 1. Prevalence of food additives across study participants.**

**Supplementary Table 2. Non-negative matrix factorization (NMF) loadings for dietary additive mixtures. Each row represents a food additive (identified by E-code and name), and each column represents an identified additive mixture pattern. Values indicate the loading coefficients (contribution weights) of each additive to each mixture, with higher values indicating stronger association with that mixture pattern. Only additives with loadings ≥0.05 in at least one mixture are included.**

**Supplementary Table 3. Food additive and mixture associations with microbial predicted pathways from PICRUSt2.**

**Supplementary Table 4. BIRDMAn effect size estimates and ASV sequences for taxa associated with dietary and additive variables.**

**Supplementary File 1. Regression modeling outputs for gut microbiota Shannon diversity analyses.**

**Supplementary File 2. XGBoost modeling results for classification and regression tasks.**
